# Child nutrition, neurodevelopment and fecal microbiota in children aged 24-60 months old in Madagascar: results from the AFRIBIOTA cross-sectional study

**DOI:** 10.1101/2025.01.16.25320667

**Authors:** Jeanne Tamarelle, Maria V. Doria, Valérie Rambolamanana, Tatamo Rajaonarivo, Ana Sousa Ferreira, Maheninasy Rakotondrainipiana, Rindra Vatosoa Randremanana, Philippe Sansonetti, Pascale Vonaesch, the Afribiota investigators

**Affiliations:** Department of Fundamental Microbiology, University of Lausanne 1015 Lausanne, Switzerland; Institut Pasteur de Paris 25-28 rue du Docteur Roux, Paris, France; Unité d’Epidémiologie et de Recherche Clinique, Institut Pasteur de Madagascar BP 1274, Ambatofotsikely, 101 Antananarivo, Madagascar; Faculdade de Psicologia, Universidade de Lisboa Alameda da Universidade, 1649-013 Lisboa, Portugal Business Research Unit (BRU-IUL), Instituto Universitário de Lisboa (ISCTE-IUL) Lisboa, Portugal; Unité de Pathogénie Microbienne, Institut Pasteur 251Z28 Rue du Dr Roux, Paris, France

**Author notes:** Contributed equally (Jeanne Tamarelle, Maria V. Doria).

## Abstract

**Introduction:** Child stunting is still a major concern worldwide with 148 million under-5 children affected in 2022. Stunting is likely to affect brain development and prevent children from reaching their full potential. This study aimed at evaluating the contribution of stunting, the fecal microbiota and other related factors in brain development in children from Madagascar.

**Methods:** Severely stunted, moderately stunted and non-stunted 2-5 years old children from the AFRIBIOTA cross-sectional study in Madagascar were submitted to the Ages and Stages Questionnaire version III, covering 5 developmental domains (communication, personal-social, problem-solving, fine motor and gross motor). Fecal samples were used for 16S rRNA gene amplicon sequencing for fecal microbiota characterization. A Structural Equation Modelling (SEM) approach was used to evaluate statistical associations, including latent variables, with direct and indirect effects.

**Results:** In all models, stunting was negatively associated with neurodevelopment, as well as low socioeconomic status. β-diversity of the microbiota was neither directly nor indirectly associated with cognitive performance but α-diversity was, in one of the tested models. Socioeconomic status, branched-chain amino acids and hemoglobin levels were associated with stunting in the SEM models tested.

**Conclusion:** Neurodevelopment was associated in this cross-sectional study with socioeconomic status and stunting. The gut microbiota α- and β-diversity were not associated with neurodevelopmental score, except for the Shannon diversity index in the complex SEM model tested. In the future, longitudinal studies assessing not only taxonomic composition but also the functional potential of the microbiome at different timepoints throughout the first years of life could shed better light on a possible, maybe also transient, role of the microbiome and metabolites released thereof on neurodevelopment.

**KEY MESSAGES:**

- What is already known on this topic? Epidemiologically, there is a well-established association between childhood undernutrition and neurodevelopmental delay. However, the etiology is unclear and there is a lack of studies proposing a comprehensible framework of stunting, neurodevelopment and other associated factors, including the intestinal microbiome.
- What does this study add? Using a SEM approach and path analysis to assess for both direct and indirect effects, we study the contribution of different factors, including the intestinal microbiota, pathogens and parasites, markers of environmental enteric disorder (EED)/intestinal inflammation, nutritional status and socioeconomic factors on child neurodevelopment. We show that branched chain amino acids (BCAAs), hemoglobin and socioeconomic status are directly associated with neurodevelopment.
- How might this study affect research, practice or policy? Our study underscores the importance of addressing iron deficiency as well as decreased levels of BCAAs in the treatment of undernourished children as they are directly associated with neurodevelopment delay. The study sets a baseline for further longitudinal studies assessing for the role of the microbiota in neurodevelopment delay associated with early life undernutrition and EED.

## INTRODUCTION

Stunting is one of the two forms of childhood undernutrition, and is defined as being too short in height for one’s age with a cutoff of two standard deviations from the median of the WHO reference [1]. Stunting is thus reflecting chronic rather than acute undernutrition. The latest UNICEF/WHO/World Bank Joint Child Malnutrition Estimates report states that in 2022 there were 148 million under-5 children considered as stunted worldwide, despite a small improvement since 2000 [1]. Evidence has accumulated on the impact of stunting in early childhood on brain development (reviewed in [2]). Prenatal and early childhood stunting can affect motor and neurodevelopment in the short-term, with effects visible as soon as at 2 years of age [3] but being maintained during the first 5 years of life [4], late childhood [5] and even into adulthood [6]. This in turn prevents children from reaching their full developmental potential, with effects such as late school enrolment and reduced educational achievements [7–10] and subsequent income losses [11]. A study by Lu *et al.* (2016) estimated that, in 2010, there were still 249 million under-5 children at risk of poor development in low-and-middle income countries (LMICs) [12]. Other factors are hindering children’s neurodevelopment, such as diet [13], male gender, poverty, rural residence, lack of cognitive stimulation [14], and anemia/iron deficiency (reviewed in [15]).

The etiology of stunting is complex and context-dependent, though there are some factors that seem to be commonly found as contributing to this syndrome, including maternal factors (nutrition, education, pregnancies), birth weight or sanitation [16, 17]. Madagascar is one of the countries which is most affected by child stunting, with a national-level under-5 stunting prevalence of about 50% [18]. Previous studies carried out in Madagascar have shown that low maternal and paternal heights [19–23], low maternal education level [23], unspaced pregnancies [20], low socioeconomic score [20], low birth weight [19, 20, 23], helminth infection [20], low food diversity score [24] and anemia [23, 25] are all associated with stunted child growth.

Enteric infections and diarrhea are both contributing to and resulting from undernutrition [26]. Diarrhea is also associated with impaired cognitive development [27] and school performance [28]. Research within the context of the AFRIBIOTA study [29] has highlighted the role of the microbiome in favoring and maintaining chronic undernutrition [30, 31], thus hindering the effectiveness of food supplementation interventions, and has shown a high prevalence of intestinal parasites [32] and enteropathogens [33] in the group of children studied. However, disentangling the respective roles of enteropathogens, the microbiome, and stunting in child development remains challenging.

Here, our objective was to study the association between stunting, neurodevelopment and the fecal microbiota in a cohort of 349 children aged 24-60 months in Madagascar. To control for potential confounders and assess for both direct and indirect effects, we used a structural equation modelling (SEM) approach. We focused our research on children older than 24 months, as previous research has shown that 70% of the deficit accumulated in height was due to faltering during the first two years of life [34]. In addition, the effects of chronic undernutrition on brain development might accumulate over time with stronger associations as children age [35].

## METHODS

### Study design

The AFRIBIOTA study is a case-control cross-sectional study carried out between December 2016 and May 2018 in Madagascar and the Central African Republic, on stunted and non-stunted children aged 24 to 60 months. The study protocol has been published previously [29]. In Madagascar, children were submitted to an additional procedure to evaluate their neurocognitive development. Briefly, children were recruited based on the following inclusion criteria: HIV-negative, not suffering from acute undernutrition (wasting) or any other severe disease, living in two neighborhoods of Antananarivo (Ankasina or Andranomanalina Isotry). Included children were admitted to the hospital for sample collection and anthropometric measurements. Stunted and control children were matched according to age, gender, neighborhood, and season of inclusion (dry or wet season).

### Measurements

The stunting status of children was assessed using the Height-for-Age z-score (HAZ), with children being considered stunted if below 2 standard deviations of the reference distribution established by the WHO [36], and considered severely stunted if below 3 standard deviations. Height was measured by trained personnel to the nearest 0.1 cm in a standing position using collapsible height boards (ShorrBoard Measuring Board, Maryland, USA). Weight was measured to the nearest 100 g using a weighing scale (KERN, ref. MGB 150K100 and EKS, Inter-équipement Madagascar).

Neurocognitive development was assessed using a translated and adapted version of the Age and Stages Questionnaires (ASQ) screening test [37, 38] in its third edition [39]. It consists of a questionnaire for each of the defined intervals for the following ages: 24, 27, 30, 33, 36, 42, 48, 54, and 60 months old, containing questions about simple tasks performed by the child in each of the following 5 areas: communication, fine motor (FM), gross motor (GM), personal-social (PS), problem-solving (PES). Trained psychologists administered the test by both observing the performance of the child on several tasks and by collecting the answers of the parents [40]. An overall development score was calculated as the sum of all five domains. No cut-offs for defining on-track development or delay were used, as these have not been validated for Malagasy children. The ASQ-3 test was carried out in a separate visit, 12 days before the main visit on average (range [-47,53]).

Biological markers were measured on the blood collected, as previously described [41]: ferritin was measured and corrected for systemic inflammation [42], using a correction factor of 0.67 if the C-reactive protein (CRP) level was above 6 mg/L. A low ferritin level was defined as below 12 μg/L. Hemoglobin values were adjusted for altitude [43] and anemia was defined as a hemoglobin level below 110 g/L [25]. The following branched-chain amino acids (BCAA) were also measured as described previously [41]: Alanine, Citrulline, Valine, Leucine and Isoleucine. Citrulline was considered low if below 7 μmol/L and elevated if above 43 μmol/L. CRP level was used as an inflammation marker, and considered elevated if above 10 mg/L. Bacterial pathogen load was determined using quantitative PCR on a set of bacterial virulence genes (*ompC, ipaH, estla, eltB, eae, bfpA, aggR, aaiC, cadF, ctxA*) as published previously [33]. Presence of a given gene was based on a Ct value < 37, and the total number of genes present in each fecal sample was reported. Helminths (*Ascaris*, *Trichuris*, *Enterobius* and *Hymenolepis*) and Protozoa (*Giardia*, *Blastocystis*) were detected using the Merthiolate-Iodine-Formaldehyde technique and the Kato-Katz technique as published previously [32].

Household/family, maternal and child additional variables were collected using a standardized paper questionnaire in French, translated to Malagasy by the staff. A socioeconomic score was built with the following categories [23]: 1) lowest socioeconomic score: no telephone or house floor made of pounded earth; 2) middle socioeconomic score: telephone, wooden or concrete house floor, no internal shower or internal kitchen; 3) highest score: telephone, wooden or concrete house floor, either an internal shower or internal kitchen (separate room). We calculated a Dietary Diversity Score (DDS) consisting of a score ranging from 0 to 7 defined as the sum of the number of food groups consumed by the child in the last 24 hours [44, 45]. The 7 different food groups were: 1) grains, roots, tubers and plantains, 2) pulses (beans, peas, lentils), nuts and seeds, 3) dairy products (milk, infant formula, yogurt, cheese), 4) flesh foods (meat, fish, poultry, organ meats), 5) eggs, 6) vitamin-A rich fruits and vegetables; and 7) other fruits and vegetables. A Low Dietary Diversity (LDD) was defined as having a DDS strictly below 4. We also included a variable on animal source food consumed by the child in the last 24 hours, including eggs, meat, fish, dairy products.

### Microbiome analyses

As described previously [31], fecal samples were collected, aliquoted on site and directly snap-frozen in liquid nitrogen and then transferred to a −80 °C freezer. DNA extraction was performed using commercial kits (QiaAmp cador Pathogen Mini or cador Pathogen 96 QIAcube HT Kit; Qiagen) following the manufacturer’s recommendations, with an additional bead-beating step. Library preparation and sequencing were carried out by Microbiome Insights. The primers used for library preparation were v4.SA501–v4.SA508 and v4.SA701–v4.SA712, as recommended by Kozich *et al.* [46]. Sequencing was then carried out on an Illumina’s MiSeq platform, using the MiSEq 500 Cycle V2 Reagent Kit (250 × 2). Demultiplexed reads were processed using the dada2 pipeline [47] and taxonomy was assigned using the Silva Reference Database (version 128).

### Statistical analysis

All analyses were carried out on R Statistical Software (version 4.1.2, R Core Team 2021) [48]. Detailed procedures, tests and model specifications are given in **SUPPL FILE 1**. We first investigated the association between nutritional status and neurodevelopment scores for each development domain and overall, using linear regression models, adjusting on confounding variables. The associations between household characteristics, maternal factors, socioeconomic status, biological markers, parasitic infections and dietary factors and i) stunting status or ii) development scores were also computed. For the latter, variables statistically significant in univariate analyses were used to build multivariate linear regression models, excluding collinear or redundant variables.

Second, we investigated the association between the fecal microbiome and neurodevelopment domains and overall neurodevelopmental score, using several data reduction approaches (α-diversity, β-diversity using Principal Coordinate Analysis (PCoA) or Hierarchical Clustering). Correlations between the number of reads for each family and neurodevelopment scores per domain and overall were computed, and a DESeq2 analysis was carried out keeping the lowest and highest quartiles of the distribution of the neurodevelopment overall score.

Finally, we built a theoretical framework of the relationships between stunting, fecal microbiota composition, neurodevelopment and other related dimensions before applying SEM and path analyses. The main outcome was child neurodevelopment. The different dimensions contributing to child neurodevelopment were grouped by blocks: socio-economic status and maternal factors, anemia, stunting, age, birth size, microbiome, BCAA and inflammation. In each of these blocks, several variables could be considered. We investigated several alternative models represented in **SUPPL FIGURE 1**:

i. The simplest SEM possible including only socioeconomic status, stunting, fecal microbiota composition and neurodevelopment, without indirect effects;
ii. A simple SEM including only socioeconomic status, stunting, fecal microbiota composition and neurodevelopment, allowing for a direct and indirect effects of socioeconomic status on neurodevelopment (through stunting and through the fecal microbiota);
iii. A complex SEM including socioeconomic status, stunting, fecal microbiota composition, neurodevelopment and other related dimensions listed above, allowing for direct and indirect effects;
iv. A complex path model including socioeconomic variables, stunting, fecal microbiota composition, neurodevelopment and other related dimensions listed above, allowing for direct and indirect effects, but without latent constructs (*i.e.*, using only observed variables).

For i), ii) and iii), we first determined latent constructs for the measurement model by calculating correlations among variables and applying confirmatory factor analysis implemented in the *lavaan* R package [49]. For microbiome data, given the high dimensionality of the data, we applied 3 different data reduction approaches to include the relevant variables in each of the four models: 1) first principal component of the PCoA, 2) α-diversity Shannon index, 3) clusters resulting from a hierarchical clustering. We then applied SEM using the *sem* function. This function was also used for path analysis (iv), without specifying a measurement model.

### Patients and public involvement

Patients were not involved in the research process. Health community volunteers were involved in the recruitment stage of the study. Both patients and health community volunteers were invited to the restitution event held in Madagascar in 2019, where local health workers, hospital heads and officials were also invited.

### Ethical statement

The study protocol of AFRIBIOTA was approved by the Institutional Review Board of the Institut Pasteur (2016-06/IRB) and the National Ethical Review Boards of Madagascar (55/MSANP/CE, 19 May 2015). All participants received oral and written information about the study. The legal representatives of the children provided written consent to participate in the study. The present analysis (AfriGutBrain) was approved by the Swiss CER-VD (BASEC-ID 2023-01834). An author reflexivity statement is attached in **SUPPL FILE 2**.

## RESULTS

### Description of household, maternal and child characteristics in the AFRIBIOTA subcohort

Three hundred forty-nine (349) children were included in this analysis, including 197 normally nourished (56%), 84 moderately stunted (24%) and 68 severely stunted children (19%). Sociodemographic characteristics of the households, mothers and children are displayed in **SUPPL TABLE 1**. Sixteen percent of the mothers of included children were undernourished, with different numbers per category of child stunting (13% for normally nourished children, 16% for moderately stunted children and 27% for severely stunted children, p = 0.026). Anemia was common in our study population (23%) and correlated with stunting severity (p = 0.035). On average children were weaned at 24 months old, leaving only 8.3% of our study population currently breastfed at the time of the study. Sixty-six percent of children had been exclusively breastfed in the first 6 months of age. *Ascaris*, *Trichuris* and *Giardia* parasitic infections were common in our study population (49%, 62% and 22% respectively). Twenty-nine percent of children had a low dietary diversity, and 8.7% did not consume any animal source foods.

### Child’s stunting status and neurodevelopment

Child stunting (HAZ score) was significantly associated with neurodevelopment in both unadjusted and adjusted models (adjusted on socioeconomic score, access to running water, child’s gender, child’s age, mother’s age at first pregnancy and mother’s educational level), with a gradient according to stunting severity (**TABLE 1**). Overall development scores ranged from 120 to 300 in our study, and in our adjusted models, moderately stunted children had a score on average -10.2 points [95% CI - 17.7, -2.7] lower than normally nourished children, while severely stunted children had a score on average -18.5 points [-26.5, -10.4] lower. Communication and PS domains were not associated with stunting. The PES domain was associated only with moderate stunting. The fine motor and gross motor domains were associated with being severely stunted only. FM score ranged from 10 to 60 in our study, and severe stunting was associated with a mean difference of -6.9 [-9.8, -4.0] points compared to normal growth. GM score ranged from 18 to 60 and severe stunting was associated with a mean difference of -5.0 [-7.3, -2.8] points compared to normal growth.

**TABLE 1.**
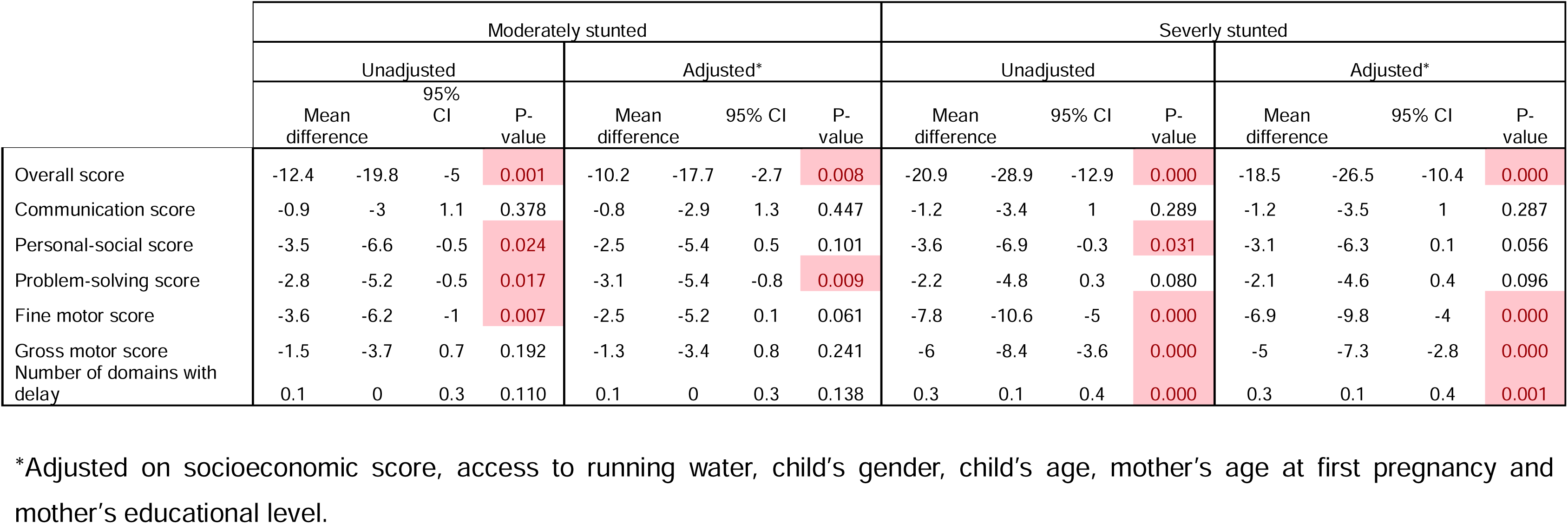
Mean difference in development score per domain and overall between normally-nourished children and moderately or verely stunted children (N = 349).

Other factors associated with neurodevelopment in each domain and with the overall neurodevelopmental score were investigated in univariate analyses (**SUPPL FIGURE 2**) and multivariate analyses (**FIGURE 1**). For the overall score and the FM score, only a higher HAZ score remained significantly associated with a higher score. In the GM domain, an older age and a higher reported birth size were associated with an increased score. In the PS domain, having access to running water in the household, drinking treated water, a higher maternal education level and a higher HAZ score were associated with an increased PS score, while an older age and having a *Blastocystis* infection were negatively associated with the PS score. Finally, in the PES domain, an older age and female sex were associated with increased score while detection of several enteropathogens’ genes in the feces was associated with a decreased score.

**FIGURE 1.**
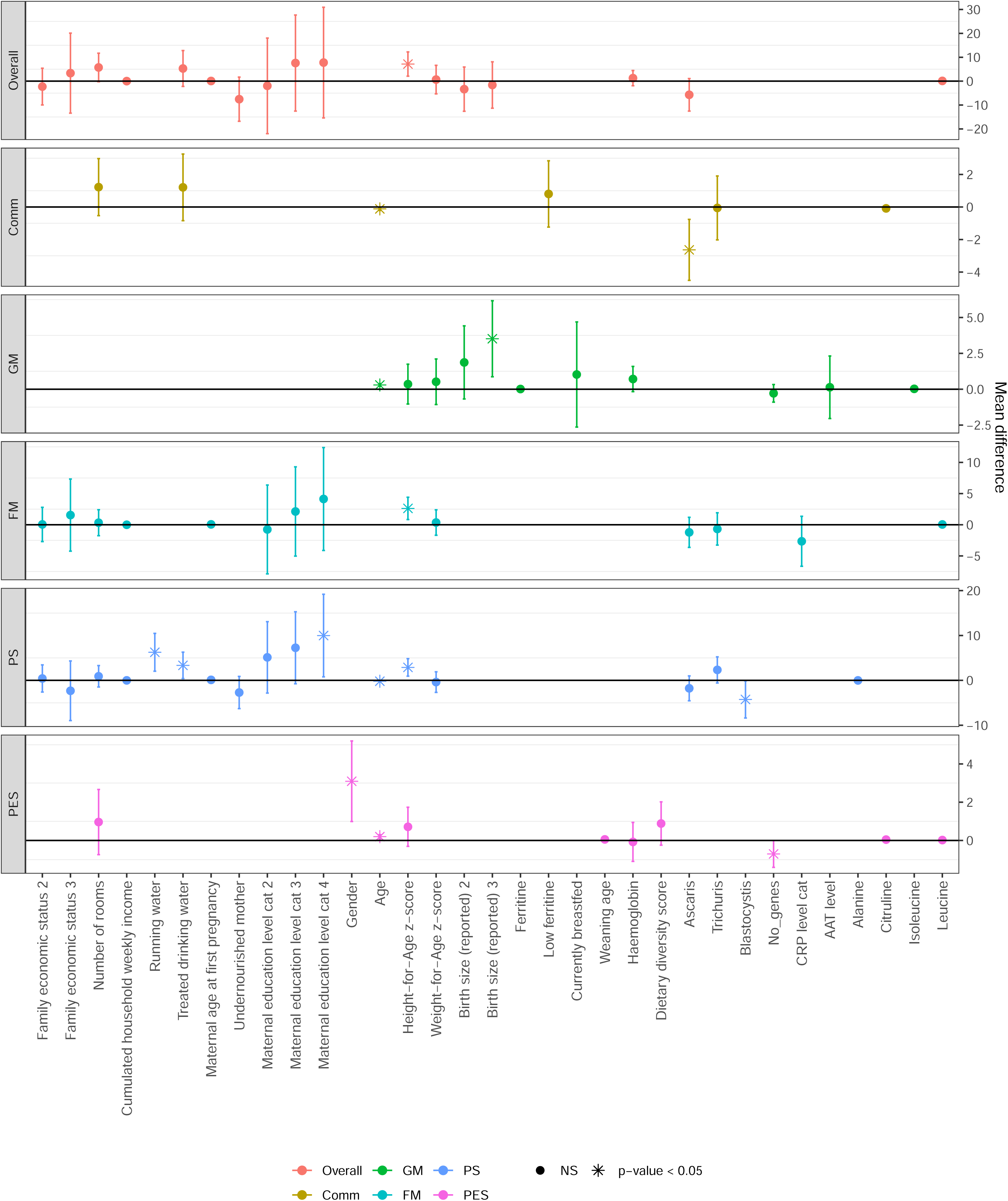
Multivariate models of association between exogenous variables and developmental scores in each domain and overall. The coding of multicategorical variables (excluding binary Yes/No variables) is as follows: Family economic status: 1) Poorest, 2) Middle, 3) Wealthiest. Maternal education level: 1) None, 2) Primary school, 3) Middle school, 4) High school or more. Birth size (reported): 1) Smaller than other babies, 2) Same size as other babies, 3) Bigger than other babies. Gender: 1) Male, 2) Female. CRP level and AAT level: 1) Normal, 2) Elevated. Reference categories are always the first level of the category (1). CRP: C-reactive protein; AAT: α-anti-trypsin; Comm: Communication; PS: Personal-Social; PES: Problem-Solving; FM: Fine Motor; GM: Gross Motor.

### Association between fecal microbiome and neurodevelopment

For subsequent analyses, fecal microbiome data was available only for 317 children. In terms of α-diversity, neither the Shannon index, nor the inverse Simpson index were associated with any of the developmental domains or with the overall neurodevelopmental score (**SUPPL FIGURE 3A**). Richness was not associated with any of the scores either (**SUPPL FIGURE 3B**). Correlations calculated between each domain score and the bacterial families revealed that one family, *Streptococcaceae*, was negatively associated with the PES score, after correcting for multiple testing (**FIGURE 2**). Further analyses with DESeq2 concerning the overall score are presented in **SUPPL FIGURE 4** and confirmed that *Streptococcaceae* was significantly associated with the PES score. The data reduction approach based on hierarchical clustering generated two microbiome clusters (**SUPPL FIGURE 5**). However, belonging to one of these two clusters did not affect the developmental scores of children (**SUPPL TABLE 3**).

**FIGURE 2.**
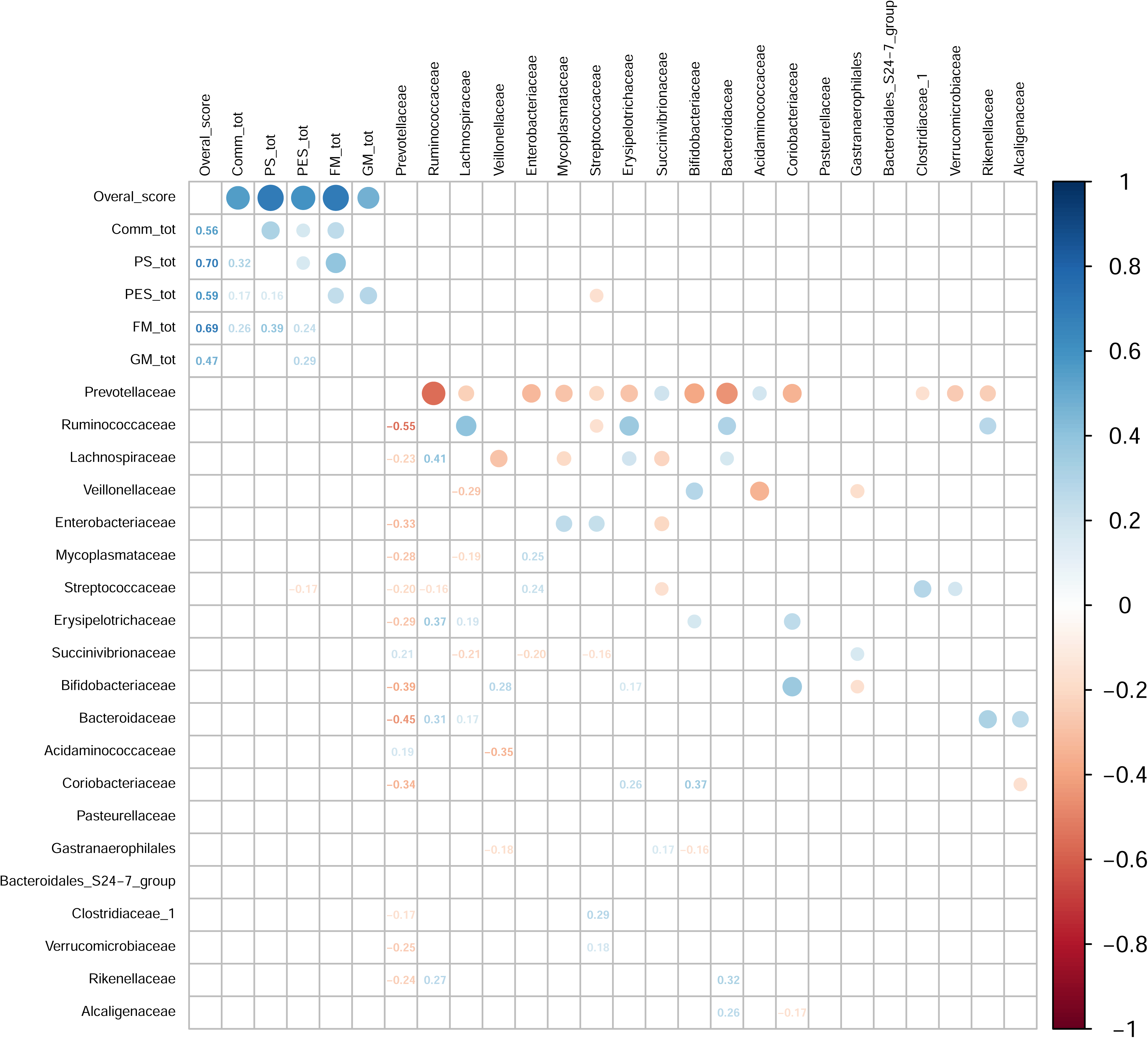
Pearson’s correlation plot between top 20 most abundant bacterial families’ relative abundance in feces and development scores in each domain and the overall neurodevelopmental score, after Benjamini-Hochberg correction for multiple testing.

### Structural equation modelling and path analysis

In SEM models, the latent variable for neurodevelopment was based on the 5 per domain scores. The socioeconomic status (SES) latent variable was based on a socioeconomic family score, maternal education level, maternal age at first pregnancy, treated water as the main source of drinking water, number of rooms in the household, as these variables were the most often associated with individual developmental domains scores (**SUPPL TABLE 2**, **SUPPL FIGURE 2**, **FIGURE 1**). Confirmatory factor analysis indicated that modelling latent constructs for neurodevelopment, SES and BCAA was fitting the data correctly (**SUPPL TABLE 4**). Therefore, these latent variables were used further for the SEM models.

Whatever the model used (simplified or complex model), stunting was always significantly associated with neurodevelopment (as a latent variable in SEMs) or the overall neurodevelopmental score (for path analysis) with a higher HAZ score being associated with a higher development score (**FIGURE 3**, **SUPPL TABLE 5**). A higher socioeconomic status was also significantly associated with a higher development score, possibly driven by the variable “number of rooms in the household” (+9.2 mean difference in overall score for each additional room in the household, p < 0.001). Age, reported birth size, maternal age at first pregnancy or α-anti-trypsin were not associated with neurodevelopment (latent variable) in multivariate SEM or the overall score in path analysis. Finally, the fecal microbiota composition, represented either by the first principal component of a PCoA or the two clusters resulting from hierarchical clustering, was never associated with neurodevelopment in the tested SEMs or with the overall score in path analysis, but the Shannon α-diversity index was, in the complex SEM model only. The standardized parameter estimates for direct mean effects for the complex SEM model (with Shannon diversity for the microbiome construct) are summarized in **FIGURE 4**.

**FIGURE 3.**
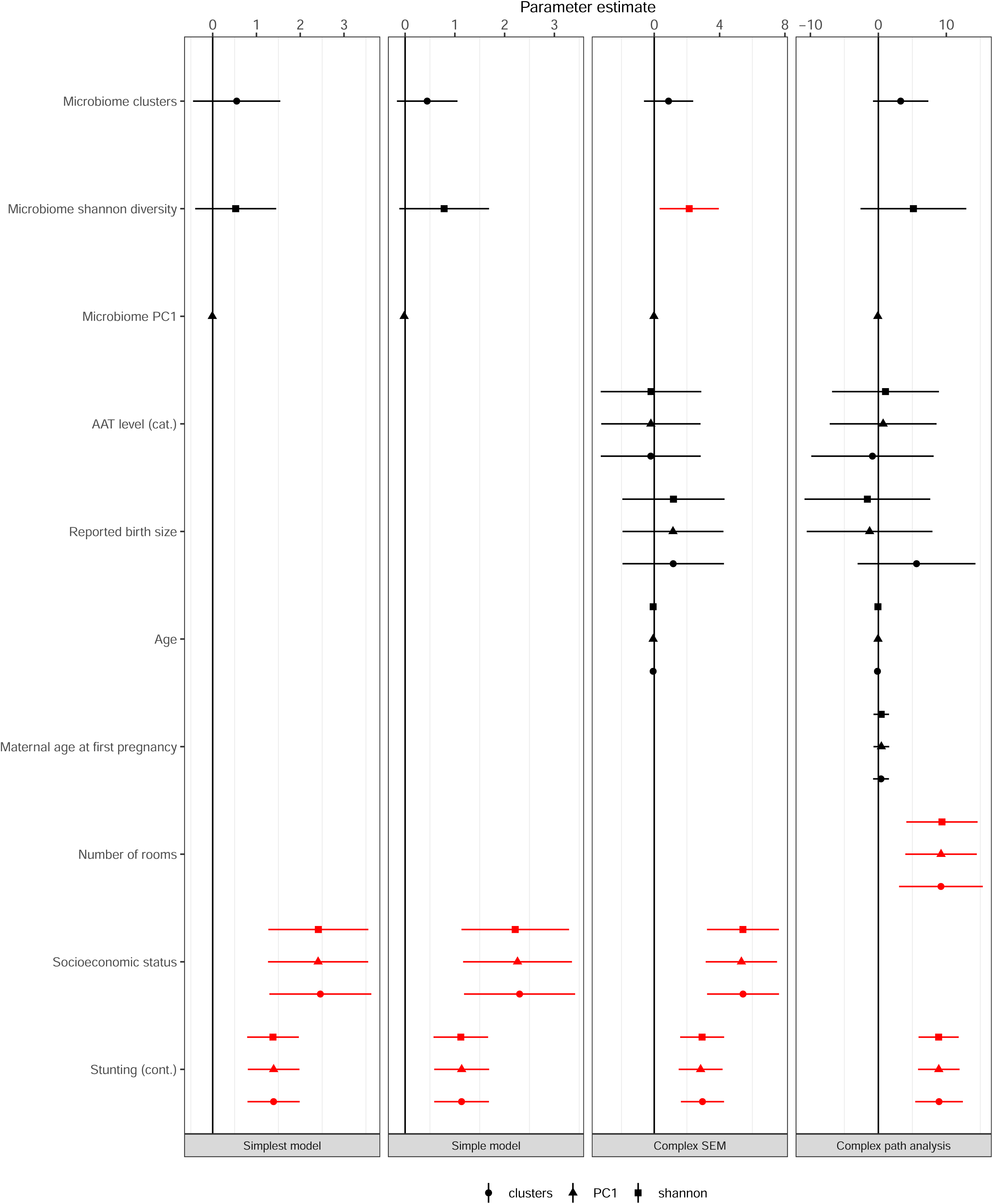
Mean direct effects of variables on neurodevelopment in the different tested models (SEM and Path analysis). Three modalities of microbiome characterization were alternatively used for each model: 1) First principal component of the PCoA, 2) Shannon U-diversity index, 3) Clusters resulting from a hierarchical clustering. Significant associations are indicated in red. PC: principal component; AAT: α-anti-trypsin.

**FIGURE 4.**
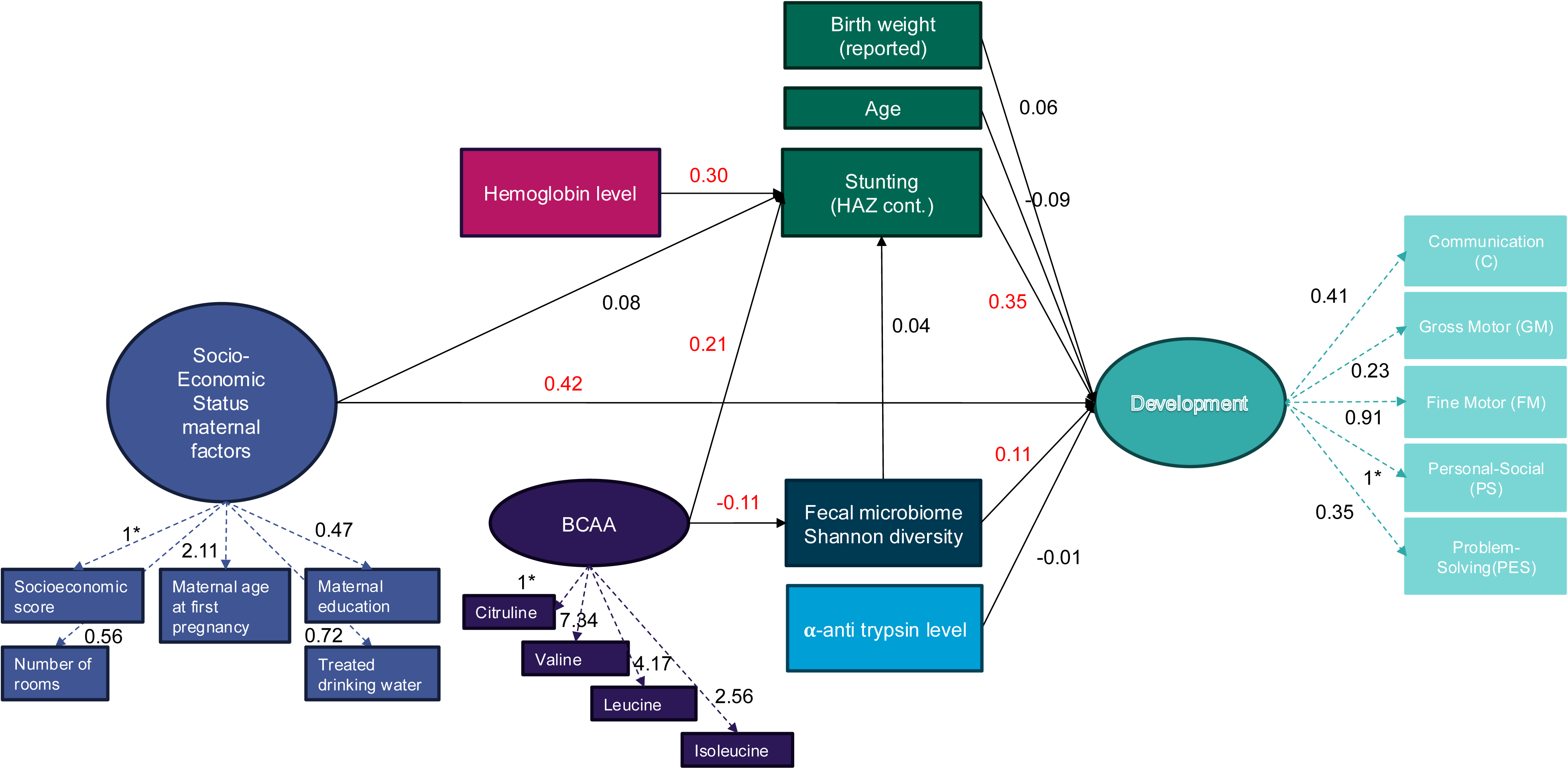
Theoretical framework including both the measurement model and the structural model, and standardized parameter estimates for direct mean effects using structural equation modelling (SEM). Significant estimates are indicated in red. Dashed arrows indicate the loadings of indicator variables in the measurement model (all significant). *Factor loading fixed to 1 for scaling purposes. HAZ: Height-for-Age Z-score; BCAA: branched-chain amico acids.

Apart from direct associations with neurodevelopment mentioned above, hemoglobin level was significantly positively associated with the HAZ score, as well as BCAA levels. And BCAA levels were negatively associated with Shannon diversity, but the magnitude of the effect was low.

Concerning indirect effects, the indirect effect of the fecal microbiota on neurodevelopment (mediated by HAZ) was always non-significant, whatever the model used (**SUPPL TABLE 6**) and the indirect effect of the socioeconomic score on neurodevelopment mediated by either the HAZ or the fecal microbiota was not significant in any of the tested models.

## DISCUSSION

In the present study, we aimed at assessing the effects of stunting and the fecal microbiota on neurodevelopment in under-5 children from Madagascar. In addition to providing a dataset on this topic from a country not yet represented in the literature, the novelty of our approach resides in two aspects. First, we sought to include information on the fecal microbiota, as it has been highlighted as a potential factor influencing neurodevelopment (reviewed in [50]). Second and more importantly, we sought to disentangle the role of stunting, the fecal microbiota and other covariates in neurodevelopment, using SEM and path analysis models, which is challenging, as the number of parameters to estimate increases drastically with the complexity of the model. Using the latent construct feature of SEM to summarize the information on the fecal microbiota (*e.g.* reducing to one latent construct the numbers of reads of 1626 bacterial amplicon sequencing variants) was not possible, as it increased the number of parameters to estimate to unrealistic targets. Instead, we first used classical data reduction approaches in the microbiome field, such as PCoA, α-diversity indices or clustering methods, and then included these constructs into the SEM models.

In this study, we confirmed the association between stunting (and stunting severity) and lower neurodevelopment scores, in a per domain approach (communication, personal-social, problem-solving, fine motor and gross motor) and a global score approach. We showed that stunting is associated with a lower overall score and with lower scores in the problem-solving, fine motor and gross motor domains, using linear regression models, adjusting on important cofactors. We further demonstrated associations between neurodevelopmental scores (overall and per domain) and reported birth size, the presence of parasites and enteropathogens, markers of socioeconomic status such as access to running water and consumption of treated drinking water, and gender. We then proposed a theoretical framework of the factors directly and indirectly influencing neurodevelopment, and we evaluated the fit of this model to our data. We used a compositional measure (latent construct) of neurodevelopment based on each of the five domains evaluated in ASQ-3 in a SEM model, as well as the overall score in a path analysis model. In these models, we showed a direct effect of stunting and low socioeconomic status on impaired neurodevelopment, and no effect of age, reported birth weight or α-anti-trypsin as a marker of overall intestinal inflammation and environmental enteric disorder. We showed a direct effect of low hemoglobin levels on stunting and therefore an indirect effect on neurodevelopment scores. High BCAA levels increased the HAZ score (and were thus negatively correlated with stunting), while they decreased the Shannon diversity. Finally, Shannon diversity alone was positively associated with increased neurodevelopment scores in the most complex SEM model.

Comparison with existing results is made difficult due to the use of different developmental scales and domain definitions in each study. The absence of effect of age on neurodevelopment was expected as the ASQ-3 tool is an age-specific test. The absence of association between neurodevelopment and reported birth size (which was well correlated with reported birth weight for children for whom we had access to this information) in the SEM models is more surprising. Of note, reported birth size was significantly associated in adjusted linear regression models only for the gross motor domain. In a previous study by Ranjitkar *et al.,* using the Bayley Scales of Infant and Toddler development, 3rd edition (BSID-III), low birth weight was associated in Nepalese children aged 6-11 months old with a lower motor score, a lower cognitive score and a lower language score [51]. This was also the case in cohorts of infants born preterm or with very low birth weights [52–55] or in meta-analysis [56] using composite scales of development. However, similar to our findings, a previous study by Alam *et al.* on children followed until 5 years of age there was no association between birth weight and cognitive development measured by the Wechsler Preschool Primary Scale of Intelligent III score, and no association between α-anti-trypsin and cognitive development either [57]. Socioeconomic status is a well-known predictor of neurodevelopment in children, although usually with small effect size, [58, 59], and usually linked to language and communication abilities. In the above-mentioned study by Alam *et al.*, socioeconomic status represented by the HME inventory and the WAMI index were positively associated with cognitive development at age 5 [57]. In another study from the MAL-ED network, socioeconomic status represented by environmental safety and healthfulness was directly positively associated with child development at 24 months old using the BSID-III evaluation tool, and indirectly associated through hemoglobin concentration [60].

Our theoretical model differed slightly from the one proposed by the MAL-ED network. We included stunting (HAZ score) in the pathway between hemoglobin level and child development, as hemoglobin level was associated with stunting in the two countries of the AFRIBIOTA project [23], leading to an indirect but positive association between hemoglobin level and child development. One of the strongest and most consistent predictors in our study was stunting (HAZ score), whatever the model tested. Stunting has been associated with neurodevelopment, mostly in the cognition and motor domains [5]. In the socioemotional domain, results are more contrasted due to poor comparability of socioemotional development measures. In longitudinal analyses, early onset of stunting and persistence until 5 years of age was particularly associated with poor development [57], which confirms the hypothesis that the effect of stunting accumulates over time, and which might be why this association is visible even in cross-sectional analyses.

Summarizing the evidence of an association between stunting and the gut microbiota in children is challenging. For instance, there are many ways to look at microbiome data, there are different study designs (cross-sectional versus longitudinal at an age where the gut microbiota still develops) and there are changes in study populations characteristics (age, geography…). Consequently, it is hard to establish whether stunting is partially caused by the microbiome or whether they both stem from the same suboptimal conditions such as infectious illness episodes, poor nutrition or poor sanitation [61]. In a recent systematic review and meta-analysis by Chibuye *et al.*, no clear signature in terms of α-diversity or β-diversity was shown between stunted and non-stunted children. In terms of bacterial taxa, no significant taxon was shared across all studies investigated, and many taxa had inconsistent associations with stunting. It seems that the two consistent traits were an abundance of pathobionts that can drive inflammation and reduce nutrient absorption, worsening the pathophysiology of stunting, and a reduction in butyrate-producers (*Faecalibacterium, Megasphera, Blautia)* and *Bifidobacterium* [61]. Hence, it was not clear to us if the gut microbiota could play a direct role on neurodevelopment or an indirect role through stunting. Our results suggest that there is no association between the high-level composition of the fecal microbiota and stunting and therefore no indirect role of the fecal microbiota on neurodevelopment, and that the direct role of the fecal microbiota on neurodevelopment is limited to α-diversity (measured by the Shannon index), with a modest effect size and not sustained in all models.

Using SEM approaches to disentangle the role of the fecal microbiota from stunting in the onset of a disease or a clinical outcome is not yet common. Some research teams have managed to include data on the gut microbiota in SEM models using different strategies. Lewis *et al.* [62] and Tokuno *et al.* [63] have defined microbiota latent variables based on some important taxa identified as significantly associated with the outcome in univariate analyses, which already represents an improvement as, to some extent, it takes into account the compositionality of microbiome data. However, this is not always doable when there are very few or even no taxa at all identified as significantly associated with the outcome, as was the case in the present study. Vu *et al.* used another strategy based on important gut biomarkers known in the literature, such as the *Enterobacteriaceae* to *Bacteroidaceae* ratio, or colonization by *C. difficile* [64]. Instead of focusing on individual taxa, we have chosen three different data reduction approaches that take into account the compositionality of microbiome data, using the Shannon α-diversity index, the first component of a PCoA, or the partition into 2 groups of a hierarchical clustering approach [65]. Our results show that our model estimates are robust to the kind of construct used for the microbiome data. The conclusions on the effect of SES, hemoglobin level, stunting, age, reported birth size or BCAA stand, whatever the microbiome variable used.

The main limits of our study reside in its small sample size and its cross-sectional design. This only allowed for moderately complex models in the SEM approach, as the number of parameters to estimate easily increases with the complexity of the model. Including several variables related to the same block was possible for some latent constructs but not all. Parasites and enteropathogens’ detection were not correlated enough to be able to build a latent construct and it was not possible to include all these variables separately in the models, as this would have increased the number of parameters to estimate to unreasonable levels. On the association between enteropathogens, illness, hemoglobin and cognitive scores, we point to the study from the MAL-ED network investigators that used a similar SEM approach [60]. Another limit resides in the fact that psychologists were not blinded to the nutritional status of the evaluated children, which could then introduce a bias in the neurodevelopment evaluation. The ability of the microbiome constructs used in the different models to capture the complexity of the fecal microbiota is limited. Indeed, the Shannon index for α-diversity is but one index, representing both the richness and evenness of taxa in our samples, but widely different microbiota compositions can have similar diversity indices. The first component of a PCoA based on Euclidean distances matrix only explained 18% of the variance observed in our microbiota data. Finally, clustering methods are subject to arbitrary choices regarding the taxonomic level, the distance metrics, the clustering methods, and in the field of the gut microbiota they have yielded equivocal results [66]. In our dataset, the existence of bacterial community clusters was not visually striking. In addition, the absence of association between β-diversity measures and neurodevelopment is further to be expected given the cross-sectional nature of our study. The use of SEM models could be greatly improved by longitudinal information, to infer causality links between variables.

Nevertheless, our study has several strengths: To date, there are almost no studies that performed SEM analysis to disentangle different factors that could contribute to a disease outcome, which is important especially in syndromes such as stunting, that are multifaceted. Further, variables included in our models had very low levels of missing data, except for birth weight, which was rarely precisely recalled by the mothers. Instead, we used reported birth size as recalled by the mothers as a proxy for birth weight. Finally, we provide data on a region where there is only scarce data to date on both early childhood neurodevelopment and the microbiome.

While future studies should focus on longitudinal approaches, our study contributes important first information on the relationship between stunting, the microbiome and neurodevelopment.

## CONCLUSION

Our study confirms the association between stunting and impaired neurodevelopment using the ASQ-3 screening tool in a population of 2–5-year-old children from Madagascar, using different statistical models. We also showed a direct effect of socioeconomic status on neurodevelopment, a limited direct effect of α-diversity on neurodevelopment and no indirect effect through stunting. Further studies should focus on longitudinal designs assessing stunting and the fecal microbiota over time to reinforce causal inferences and establish a comprehensive model of stunting, the fecal microbiota and neurodevelopment in under-5 children.

## Supporting information

STROBE checklist

## Data Availability

All data produced in the present study are available upon reasonable request to the authors

## ACKNOWLEDGMENTS

We would like to thank Kurt Long for helpful discussions regarding the SEM model approach. We further would like to thank all field workers, doctors, community health workers and families implicated in Afribiota.

## List of Afribiota Investigators (in alphabetical order)

Robert Barouki, Hôpital Necker-Enfants maladies, Paris, France

Alexandra Bastaraud, Institut Pasteur de Madagascar, Antananarivo, Madagascar

Jean-Marc Collard, Institut Pasteur de Madagascar, Antananarivo, Madagascar

Maria Doria, Institut Pasteur, Paris, France

Darragh Duffy, Institut Pasteur, Paris, France

B. Brett Finlay, University of British Columbia, Vancouver, Canada

Serge Ghislain Djorie, Institut Pasteur de Bangui, Bangui, Central

African Republic Tamara Giles-Vernick, Institut Pasteur, Paris, France

Bolmbaye Privat Gondje, Complexe Pédiatrique de Bangui, Bangui, Central African Republic

Jean-Chrysostome Gody, Complexe Pédiatrique de Bangui, Bangui, Central African Republic

Milena Hasan, Institut Pasteur, France

Nathalie Kapel, Hôpital Pitié-Salpêtrière, Paris, France

Jean-Pierre Lombart, Institut Pasteur de Bangui, Bangui, Central African Republic

Synthia Nazita Nigatoloum, Complexe Pédiatrique de Bangui, Bangui, Central African Republic

Laura Wegener Parfrey, University of British Columbia, Vancouver, Canada

Maheninasy Rakotondrainipiana, Institut Pasteur de Madagascar, Antananarivo, Madagascar

Rindra Vatosoa Randremanana, Institut Pasteur de Madagascar, Antananarivo, Madagascar

Annick Robinson, Centre Hospitalier Universitaire Mère Enfant de Tsaralalana, Antananarivo, Madagascar

Pierre-Alain Rubbo, Institut Pasteur de Bangui, Bangui, République Centrafricaine

Philippe Sansonetti, Institut Pasteur, Paris, France

Laura Schaeffer, Institut Pasteur, Paris, France

Ionela Gouandjika-Vasilache, Institut Pasteur de Bangui, Bangui, République Centrafricaine

Pascale Vonaesch, Institut Pasteur, Paris, France

Sonia Sandrine Vondo, Complexe Pédiatrique de Bangui, Bangui, Central African Republic

Inès Vigan-Womas, Institut Pasteur de Madagascar, Antananarivo, Madagascar

