## Supplemental file 1 for "Child nutrition, neurodevelopment and fecal microbiota in children aged 24-60 months old in Madagascar: results from the AFRIBIOTA cross-sectional study"

**SUPPLEMENTARY FILE 1. ADDITIONAL METHODS**

1) Univariable and multivariable linear regression models for HAZ score (exposure) and neurodevelopment scores (outcome) – **TABLE 1**

Models were adjusted on socioeconomic score, access to running water, child’s gender, child’s age, mother’s age at first pregnancy and mother’s educational level.

For each model the assumptions were verified graphically using diagnostic plots (function *plot(model)* in the in the *base* library)

- Residuals vs Fitted plot for the linear relationship assumption;
- Normal Q-Q plot for the normality of the distribution of residuals;
- Scale-Location plot for the homoscedasticity of the variance of residuals;
- Residuals vs Leverage plot for identifying influential cases or extreme values.

In addition, the Breusch–Pagan test was used to check homoskedasticity in univariable and multivariable linear regression models, using the function *bptest(model)* in the *lmtest* package.

2) Univariable and multivariable linear regression models for several exposures and neurodevelopment scores (outcome) – **SUPPL TABLE 2, SUPPL FIGURE 2 and FIGURE 1**

Similarly, the assumptions for linear regression models were verified graphically using diagnostic plots, only for multivariable models due to high number of exposures and outcomes in univariable models.

3) Statistical tests for exposures associated with stunting (outcome) – **SUPPL TABLE 1**

Absolute numbers and percentages were given in each category of the outcome (normally nourished, stunted, severely stunted) for binary and categorical variables, and the Fisher’s exact test was used for statistical significance. Means (minimum-maximum) were given in each category of the outcome for continuous variables, and the Kruskal-Wallis rank sum test or the Pearson’s Chi-squared tests were used for statistical significance depending on the size of expected values.

4) Association between fecal microbiome and neurodevelopment scores

- α-diversity

α-diversity was evaluated on unaggregated reads (Amplicon Sequencing Variants level) using:

- the Shannon index and the inverse Simpson index in the *diversity* function of the *vegan* package – **SUPPL FIGURE 3A**
- The richness in the *estimate* function of the *vegan* package – **SUPPL FIGURE 3B**

Univariable linear regression models were run on each of these indices (exposures) and neurodevelopment scores (outcome) and dot plots with the regression line were plotted. The Pearson’s r correlation and the p-value were added to each plot.

Linear regression models adjusting on the age range of the ASQ-3 questionnaire and the HAZ score were also computed but are not reported as all coefficients for diversity indices are non-significant.

- Individual taxa

Associations between individual taxa and neurodevelopment scores were evaluated at the family levels on relative abundances.

- Correlations between individual bacterial families and neurodevelopment scores were calculated using Pearson’ r. The results were corrected for multiple testing using the Benjamini-Hochberg correction – **FIGURE 2**
- We also performed differential abundance testing using the package DESeq2 [1] at the family level, with each family used alternatively as the outcome and overall neurodevelopment delay as a binary covariate. Because no cut-offs adapted to Malagasy children have been proposed so far, we used the first and last quartiles of the distribution of each continuous score as a binary covariate for DESeq2. This algorithm uses absolute abundances of reads. Because many taxa had at least one zero, no estimation of size factors was possible. We thus computed a geometric mean removing zero counts. The results were also corrected for multiple testing using the Benjamini-Hochberg correction – **SUPPL FIGURE 4**
- β-diversity

β-diversity was evaluated using either Principal Coordinate Analysis (PCoA) or Ascending Agglomerative Hierarchical Clustering approaches.

- For PCoA, distances between samples were calculated using alternatively i) the Euclidean distance on relative abundance of reads, ii) the Bray-Curtis distance on relative abundance of reads, iii) the Euclidean distance on center-log ratio-transformed data (Aitchison distance). The difference in β-diversity between delayed and non-delayed children was tested statistically using PERMANOVA. However, because this implied using a cut-off to define delay, the results are not presented in the manuscript. None were significant, except for the FM domain with Euclidean distance and the GM domain with Aitchison distance.
- For hierarchical clustering, distances between samples were calculated using the Euclidean distance on relative abundance of reads and the Ward linkage, using the option “Ward.D2” in the *hclust* function. The number of clusters was determined visually using the elbow method [2] based on the Calinski-Harabasz index – **SUPPL FIGURE 5**

5) SEM and path analysis models – **SUPPL FIGURE 1**

- For the measurement model, latent variables were determined as follows: we first calculated pairwise correlations among variables of a block. If variables were continuous, we used Spearman’s r coefficient. If they were categorical, we used Cramer’s V statistic, and if one was continuous and the other one categorical, we used the square root of the R^2^ of an ANOVA test. By visual inspection of the correlation plots, it became obvious that only three blocks were susceptible to be used for latent constructs: neurodevelopment, socioeconomic status and maternal factors and BCAA. All the other blocks with at least 4 variables had correlations too low (< 0.2) to build latent constructs. The selection of observed and latent variables to include in the measurement model was based on the following criteria: 1) at least 4 indicator variables to determine a latent construct to allow identification of the model, 2) if less than 4 indicator variables, replacement of the latent construct by a variable for which the association with endogenous variables is the strongest.
- It was not possible to include all taxa in the model to build a microbiome latent construct, therefore we used three alternative approaches:
  - using the first construct of a PCoA using Euclidean distances on relative abundance data, explaining 18% of the variance of the microbiome dataset;
  - using the clusters issued from the hierarchical clustering procedure described above;
  - using the α-diversity Shannon index.

For each model (SEM or path analysis, we tried each of these 3 constructs and reported all results) – **FIGURE 3**

- For the structural model, the main outcome was neurodevelopment, inserted in the model either as a latent variable for SEM models or as the overall neurodevelopment score for path analysis.
- The selection of variables for the structural model of SEMs and path models was based on the following parsimonious criteria: 1) previously demonstrated association with child development and/or stunting in univariate analysis, 2) quality of the collected information and low rate of missing data.
