## Supplemental file 2 for "Child nutrition, neurodevelopment and fecal microbiota in children aged 24-60 months old in Madagascar: results from the AFRIBIOTA cross-sectional study"

**SUPPLEMENTARY FILE 2. Author reflexivity statement**

1. *How does this study address local research and policy priorities?*

Malnutrition is one of the main Public Health Issues in Madagascar. As the study was further performed in some of the most disfavoured districts of Antananarivo, the project directly adresses important health issues of the populations studied.

2. *How were local researchers involved in study design?*

The local researchers, especially the senior researchers, where involved in the study design from the very beginning, contributing also to the writing of the funding applications, the ethical protocols and all Standard Operating Procedures. Several of the Work Packages of Afribiota were also led by local researchers. Regular meetings were held with all involved stakeholders to re-orient the project, discuss and overcome hurdles and later discuss results and frame the manuscripts.

3. *How has funding been used to support the local research team(s)?*

The Afribiota project has supported capacity building in both study sites. In Bangui, the project allowed a microbiologist to follow a training course at Institut Pasteur de Madagascar on microbiology and immunology. Furthermore, Prof. Vonaesch held several sessions on DNA extraction and molecular biology for the local team and also trained the local researchers in clinical data management and monitoring. Last, the project contributed essential equipment to the Institut Pasteur de Bangui, including a NanoDrop machine, a ultrasound bath as well as material for molecular biology.

In Madagascar, the project supported three PhD students: Azimdine Habib, in the meantime graduated in parasitology from the University of Antananarivo, Zo Andrianamantena, about to graduate at the University of Antananarivo in Immunology and Elliot Rakotomanana, who graduated in anthropology at the University of Bordeaux. There was also a medical student performing her medical thesis on the project. Furthermore, several members of the clinical team got trained in clinical research and good clinical practices through online courses and the main clinical study manager benefited from a MPH in Public Health at the Institut Pasteur once the recruitments ended.

Furthermore, the local researchers visted twice in Paris for steering meetings of the project and a delegation of researchers from Madagascar also visited the Bangui site. Last, there were two visits of the European team, once in Bangui and twice in Madagascar, where world-known researchers from the Scientific Advisory Board of Afribiota gave presentations on their research. The teams from Madagacar and the Central African Republic also visited Institut Pasteur for a mid-project evaluation and brainstorming session.

Finally, Prof. Pascale Vonaesch and several other lead scientists supported the local researchers in the writing of their manuscript, of which several are first and/or last authored by local researchers.

4. *How are research staff who conducted data collection acknowledged?*

The main clinical study manager as well as the psychologists performing the cognitive tets are co-authors of this article. The data collectors for general clinical data/sample collection are further acknowleged in the manuscript.

5. *How have members of the research partnership been provided with access to study data?*

All members of Afribiota had access to all data of the project. They are further encouraged to use the data generated through the project for secondary analyses. Follow-up analyses of the project are currently ongoing, of which several are initiated and performed by local researchers.

6. *How were data used to develop analytical skills within the partnership?*

Several local researchers performed their own analyses on the data gathered within the Afribiota project and published the results as lead author. In the current manuscript, seen the advanced statistical methods used, the analysis was performed by a postdoctoral fellow specialised in statistics.

7. *How have research partners collaborated in interpreting study data?*

All researchers were involved in the final data interpretation and the critical reviewing of the manuscript.

8. *How were research partners supported to develop writing skills?*

Prof. Vonaesch and several other senior researchers from the Afribiota consortium have co-written research articles with local researchers from Madagascar and the Central African Republic, thus training them in scientific writing.

9. *How will research products be shared to address local needs?*

There has been a restitution event in Madagascar in 2019, where local health workers, hospital heads and officials have been invited. Part of the results of Afribiota have been presented at this event. Institut Pasteur de Madagascar is in regular contact with the Ministery of Health and the results will thus be shared with the local health authorities.

10. *How is the leadership, contribution and ownership of this work by LMIC researchers recognised within the authorship?*

The Afribiota project is co-led by three Institutions: Institut Pasteur, Paris, Institut Pasteur de Madagascar and Institut Pasteur de Bangui, Central African Republic. This article was based on data from the Madagascar site, of which Dr. Rindra Randremana was the local responsible/main principal investigator. The study was supervised by Dr. Maria Doria, a graduated psychologist and early childhood specialist, who regularly visited Madagascar to work with the two local psychologists, Valérie Rambolamanana and Tatamo Rajaonarivo. The project was jointly led by Dr. Maria Doria, Prof. Pascale Vonaesch (as co-main leader of Afribiota) and Dr. Rindra Randremanana (as main site responsible). All researchers directly involved in the work are listed as co-authors. The consortia member that facilitated the work are listed as group authorship within the consortium signature.

11. *How have early career researchers across the partnership been included within the authorship team?*

There are several early career researchers on the article: Dr. Jeanne Tamarelle is a postdoctoral fellow and Prof. Pascale Vonaesch a tenure-track Assistant Professor. Furthermore, Valérie Rambolamanana and Tatamo Rajaonarivo where recent psychology graduates at the time this study was performed and Maheninasy Rakotondrainipiana recent graduate from medical school.

12. *How has gender balance been addressed within the authorship?*

We acknowlege that there is a gender imbalance, as the first authors as well as the last authors are female on this article. Overall, 8 out of 9 authors are female. This gender imbalance is due to the fact that this Work Package of the Afribiota project was led by chance by several female researchers. In Afribiota overall, there was a gender balance between female and male, as visible in the Afribiota investigators, where 12 out of 29 researchers are male. The authorship reflects the contribution of each team member to the currrent authorship based on international authorshiop guidelines.

13. *How has the project contributed to training of LMIC researchers?*

The project has trained two recent psychology graduates, Valérie Rambolamanana and Tatamo Rajaonarivo, in clinical research and in the use of early childhood development tools. This training was provided over several years through several in-person trainings by Dr. Doria on site in Madagascar as well as bimonthly meetings of Dr. Doria with the Malagasy psychologists to discuss hurdles in the field, data recording and analysis. Furthermore, through Afribiota, Maheninasy Rakotondrainipiana has been trained in clinical research. She also benefited from online classes in good clinical practices as well as a Master’s in Public Health at Institut Pasteur, in which she was enrolled after the end of the recruitments.

14. *How has the project contributed to improvements in local infrastructure?*

The Afribiota project was hosted at Institut Pasteur de Madagascar. No special infrastructure was needed for the present project. However, through Afribiota, several new tools were brought to Madagascar, including a Malagasy translation of the Ages and Stages Questionnaire, that was used in this study.

15. *What safeguarding procedures were used to protect local study participants and researchers?*

All data was recorded on coded computers and paper questionnaires were stored in a closed cupboard at the Institut Pasteur de Madagascar. All participant data is coded, *i.e.* given a special identifier that does not contain the name of the participant. The correspondance list is stored in a closed cupboard at Institut Pasteur de Madagascar. A charter for the Afribiota project is protecting the data ownership by all three participating Institutions, saveguarding preferential access to the data to the local researchers from the implicated Institutions while allowing all scientist to request access to the data with a project plan.
