## Supplemental file 3 for "Child nutrition, neurodevelopment and fecal microbiota in children aged 24-60 months old in Madagascar: results from the AFRIBIOTA cross-sectional study"

**SUPPLEMENTARY FILE 3. SUPPLEMENTARY FIGURES WITH LEGENDS**

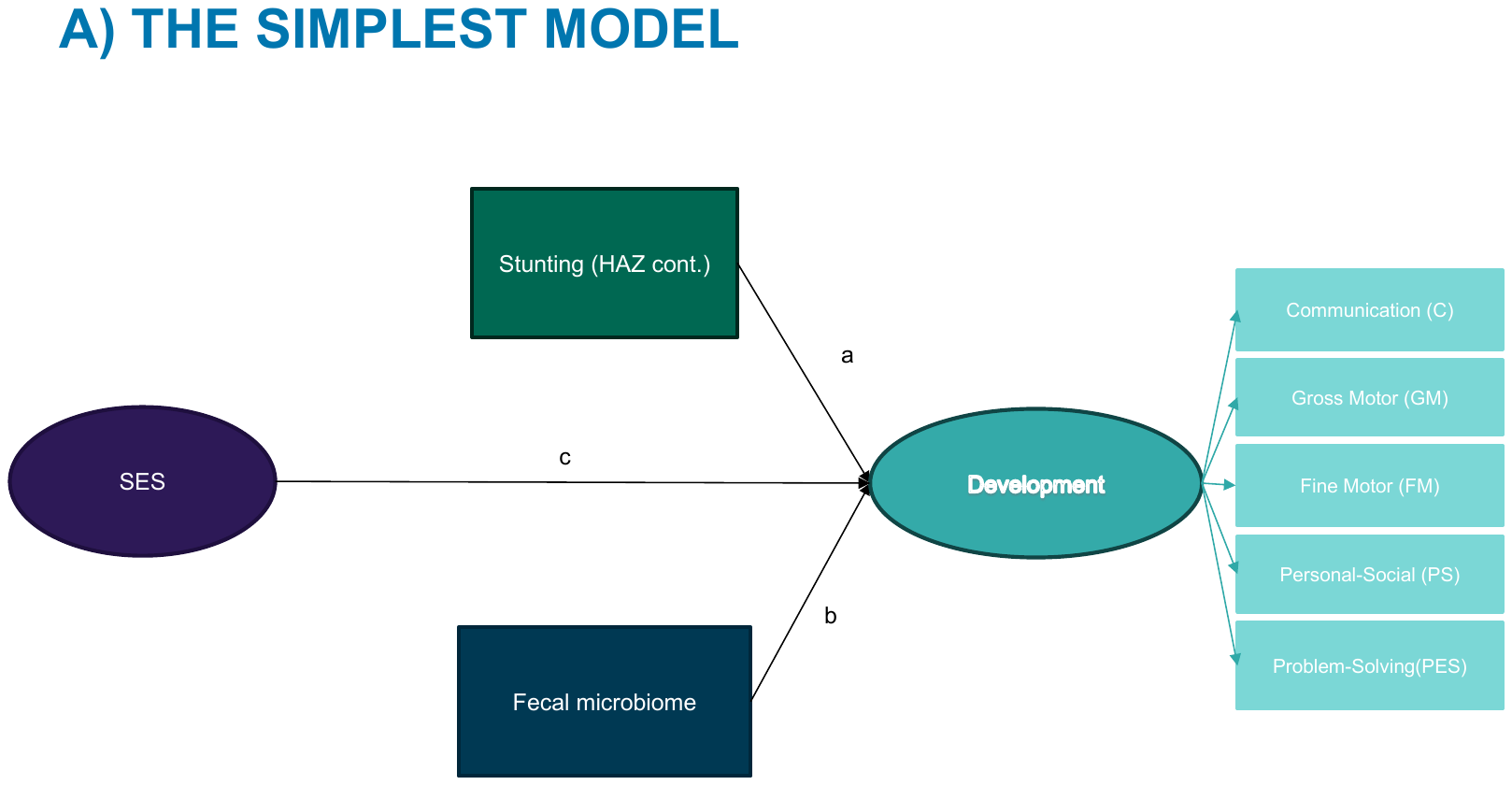

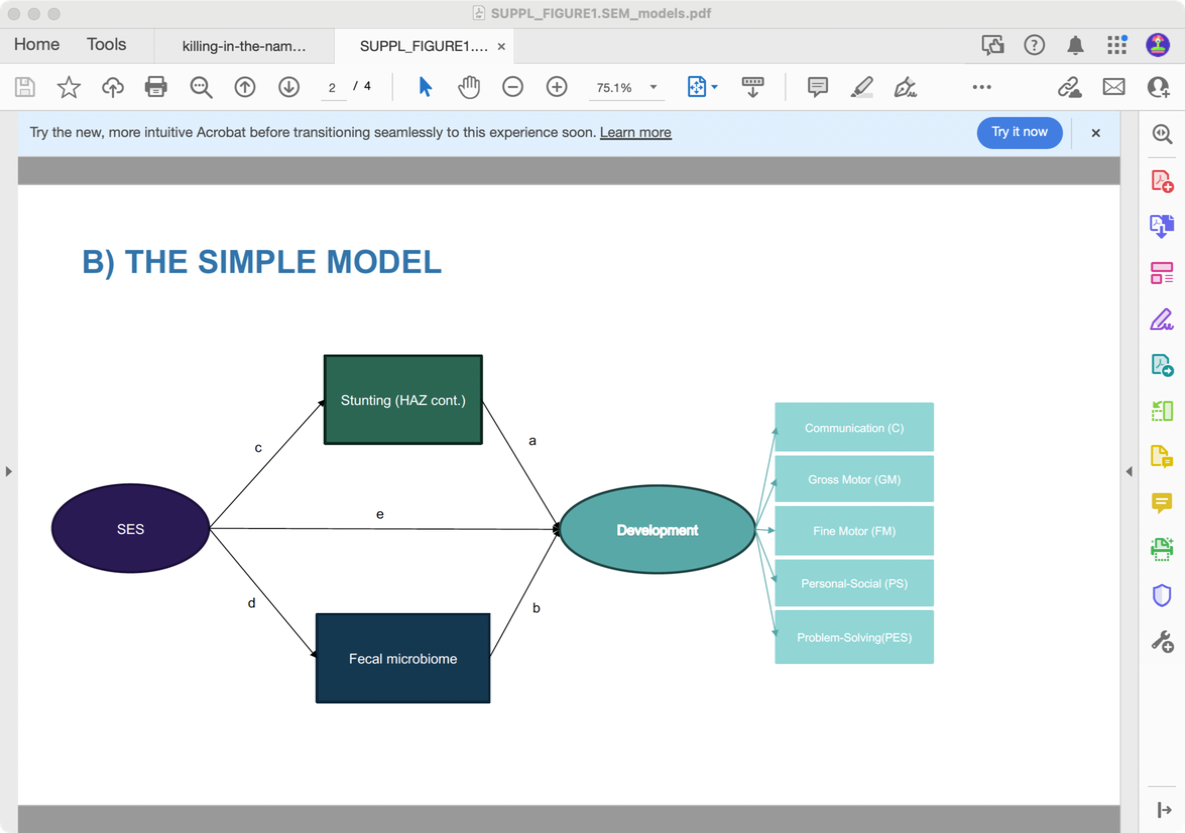

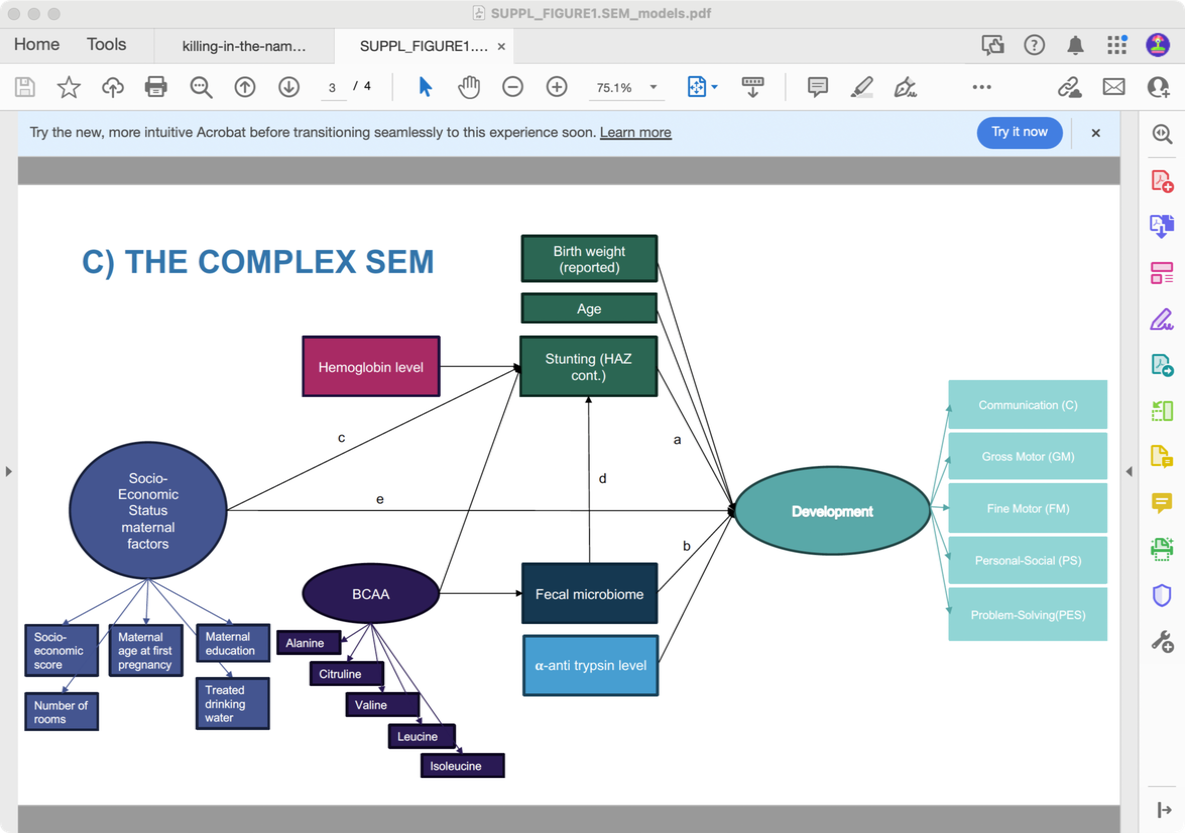

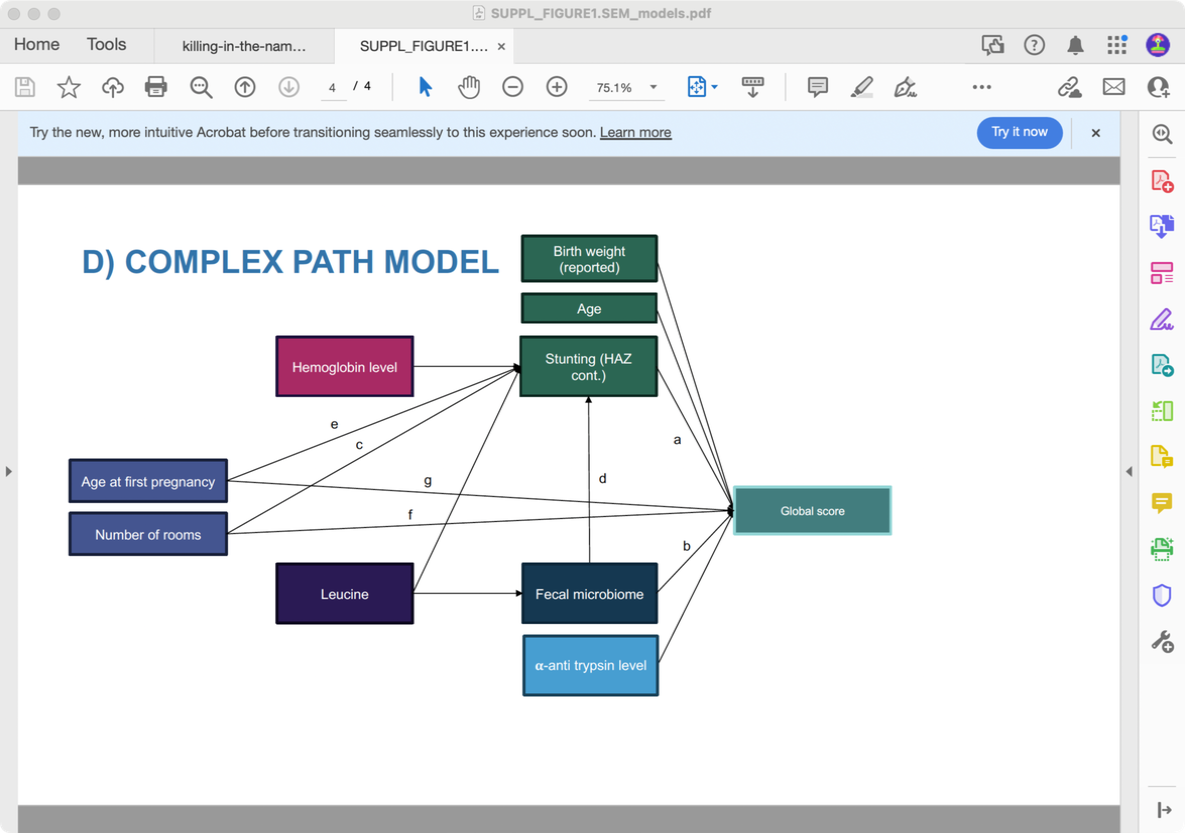

**SUPPL FIGURE 1.** Theoretical models tested with observed and indicator variables represented as squares and latent variables represented as circles. A) The simplest SEM model includes only direct effects of the socioeconomic status, the fecal microbiome and stunting on neurodevelopment; B) the simple SEM model includes direct effects of the socioeconomic status, the fecal microbiome and stunting on neurodevelopment and an indirect effect of the socioeconomic status on neurodevelopment through stunting and through the fecal microbiome; C) The complex SEM model includes the socioeconomic status, the fecal microbiome, stunting, anemia, BCAA, α-anti-trypsin level, age and reported birth size, allowing for direct and indirect effects on neurodevelopment; D) The complex path model includes only observed variables (number of rooms, maternal age at first pregnancy, the fecal microbiome, stunting, anemia, leucine, α-anti-trypsin level, age and reported birth size), without latent constructs, allowing for direct and indirect effects on the overall development score.

HAZ: Height-for-Age Z-score; BCAA: branched-chain amino acids.

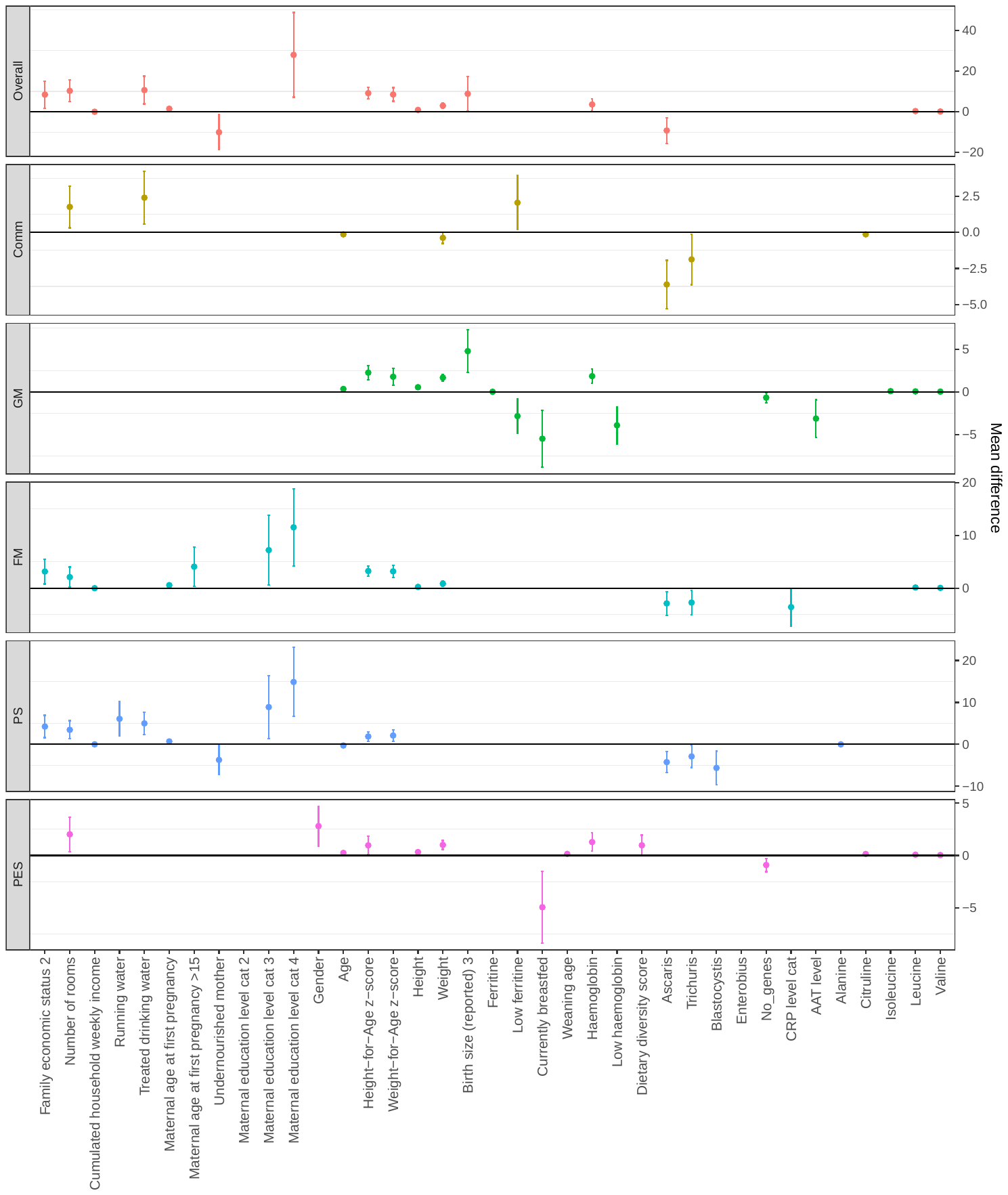
**SUPPL FIGURE 2.** Statistically significant associations between exogenous variables and developmental scores in each domain and in the overall neurodevelopmental score in univariate analyses.

The coding of multicategorical variables (excluding binary Yes/No variables) is as follows:

Family economic status: 1) Poorest, 2) Middle, 3) Wealthiest. Maternal education level: 1) None, 2) Primary school, 3) Middle school, 4) High school or more. Birth size (reported): 1) Smaller than other babies, 2) Same size as other babies, 3) Bigger than other babies. Gender: 1) Male, 2) Female. CRP level and AAT level: 1) Normal, 2) Elevated.

Reference categories are always the first level of the category (1).

CRP: C-reactive protein; AAT: α-anti-trypsin; Comm: Communication; PS: Personal-Social; PES: Problem-Solving; FM: Fine Motor; GM: Gross Motor.

A) **Evenness**

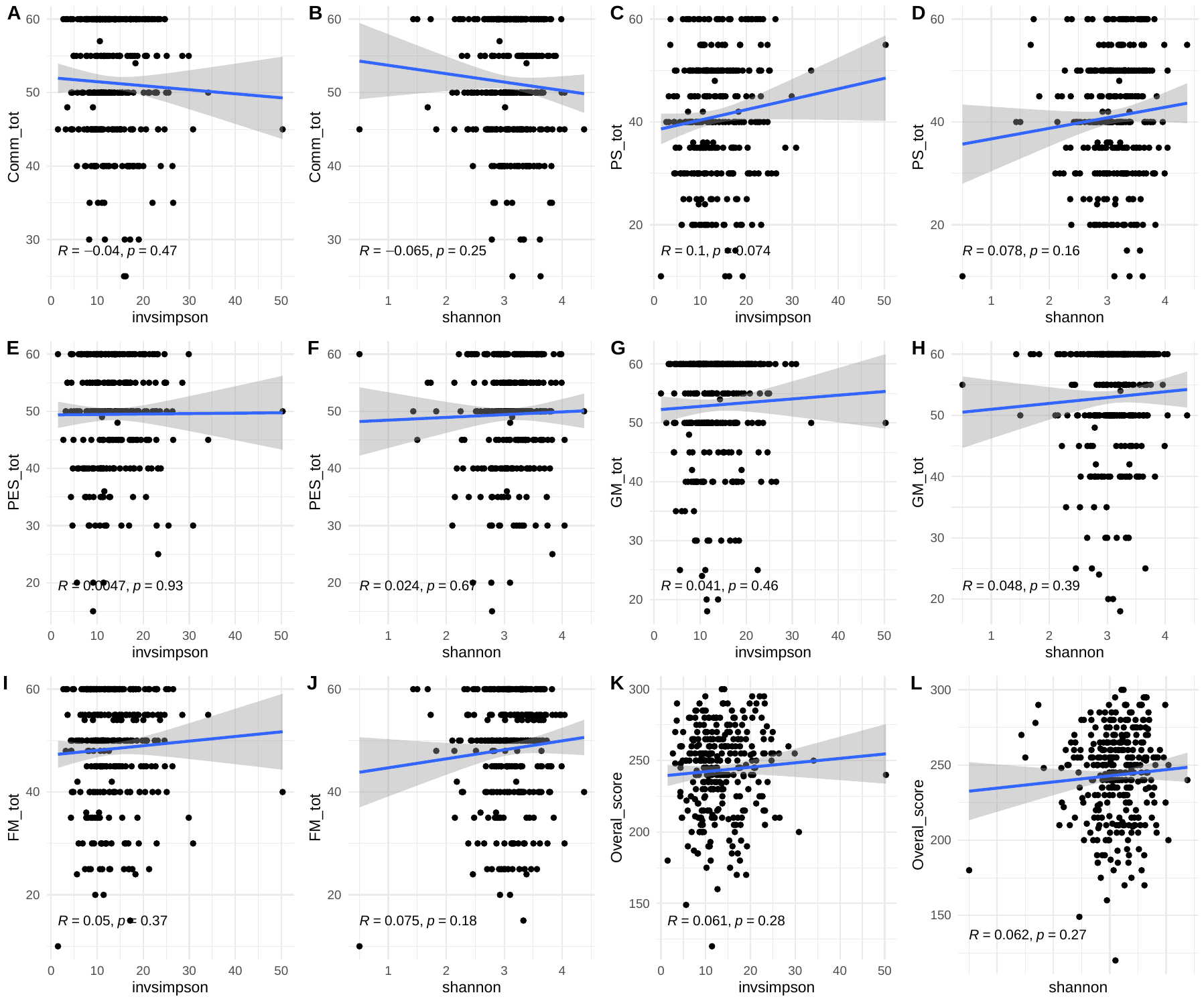

B) **Richness**

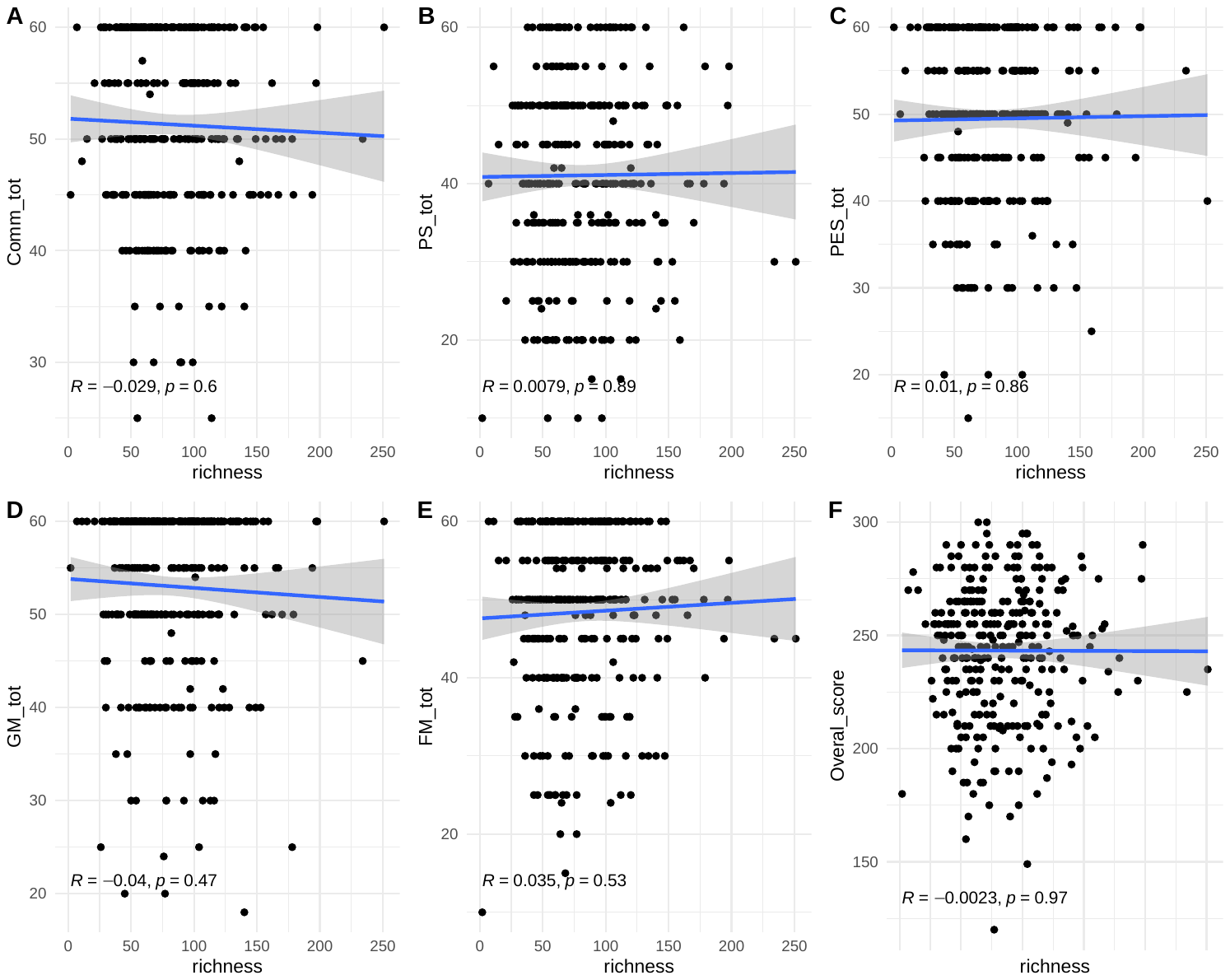

**SUPPL FIGURE 3.** Fecal microbiota composition and neurodevelopment. A) Linear regressions between alpha diversity represented by the Shannon index (A,C,E,G,I,K) or the inverse Simpson index (B,D,F,H,J,L) and development scores in each domain and in the overall neurodevelopmental score; B) Linear regressions between alpha diversity richness and development scores in each domain and in the overall neurodevelopmental score.

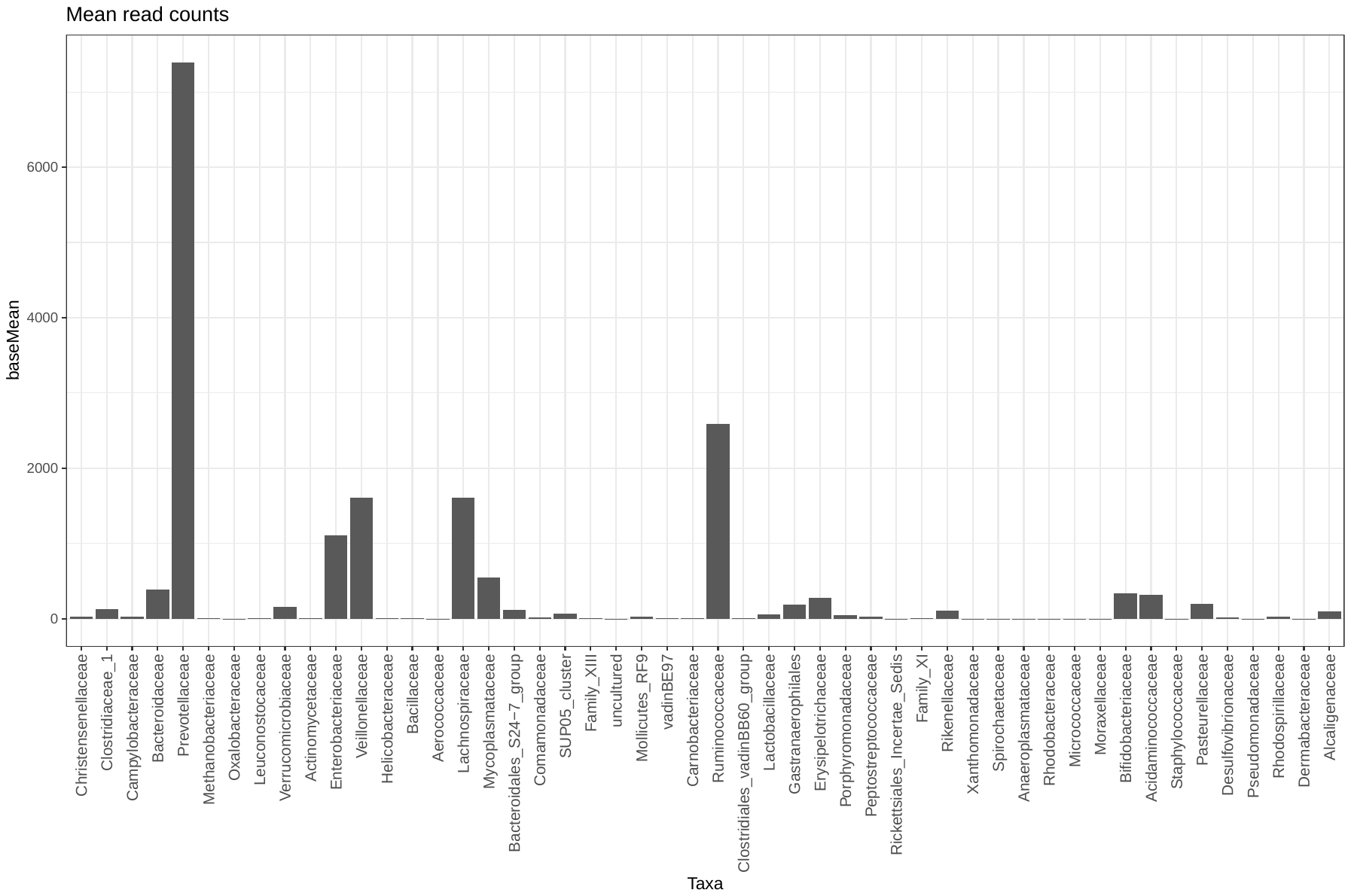

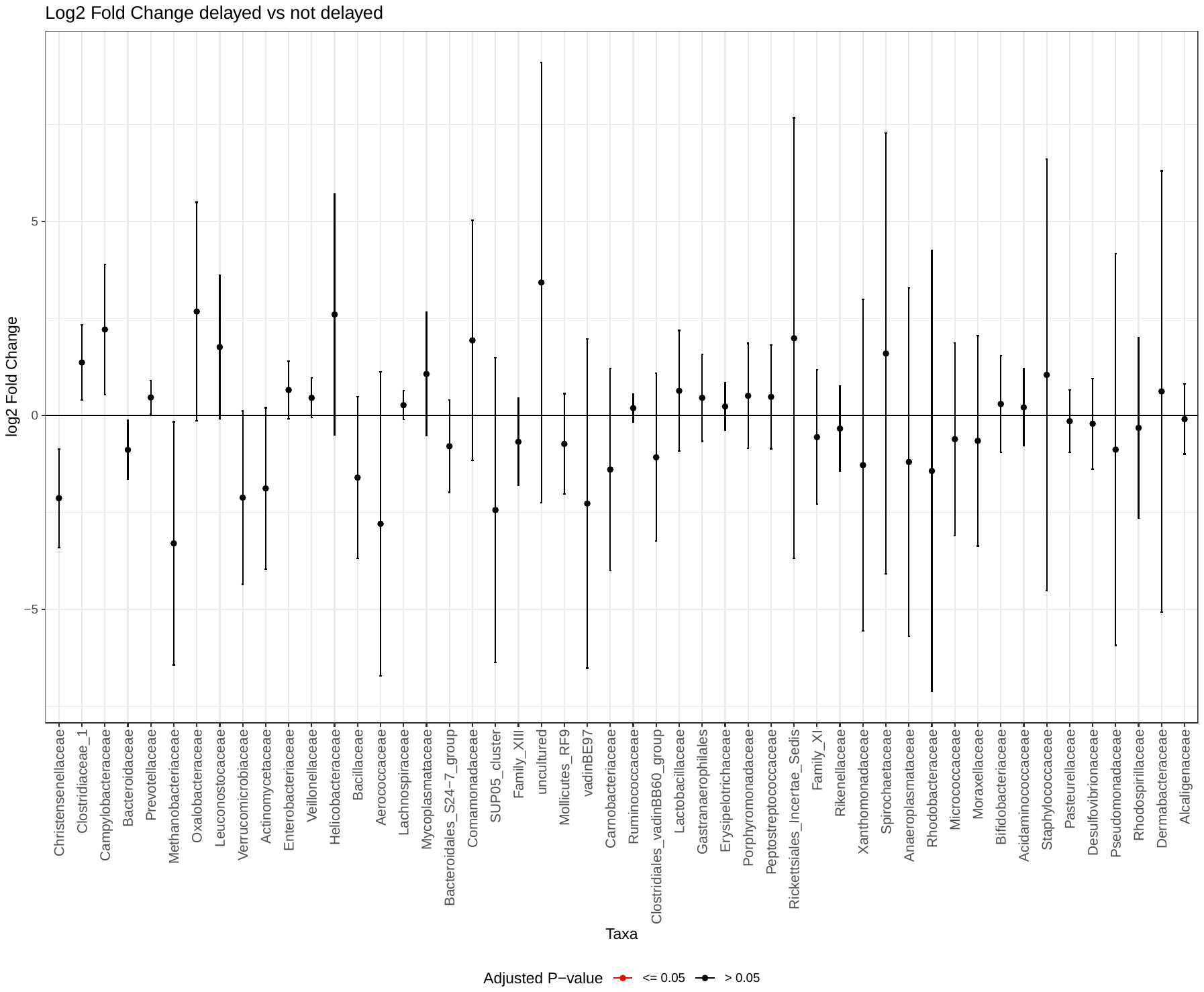

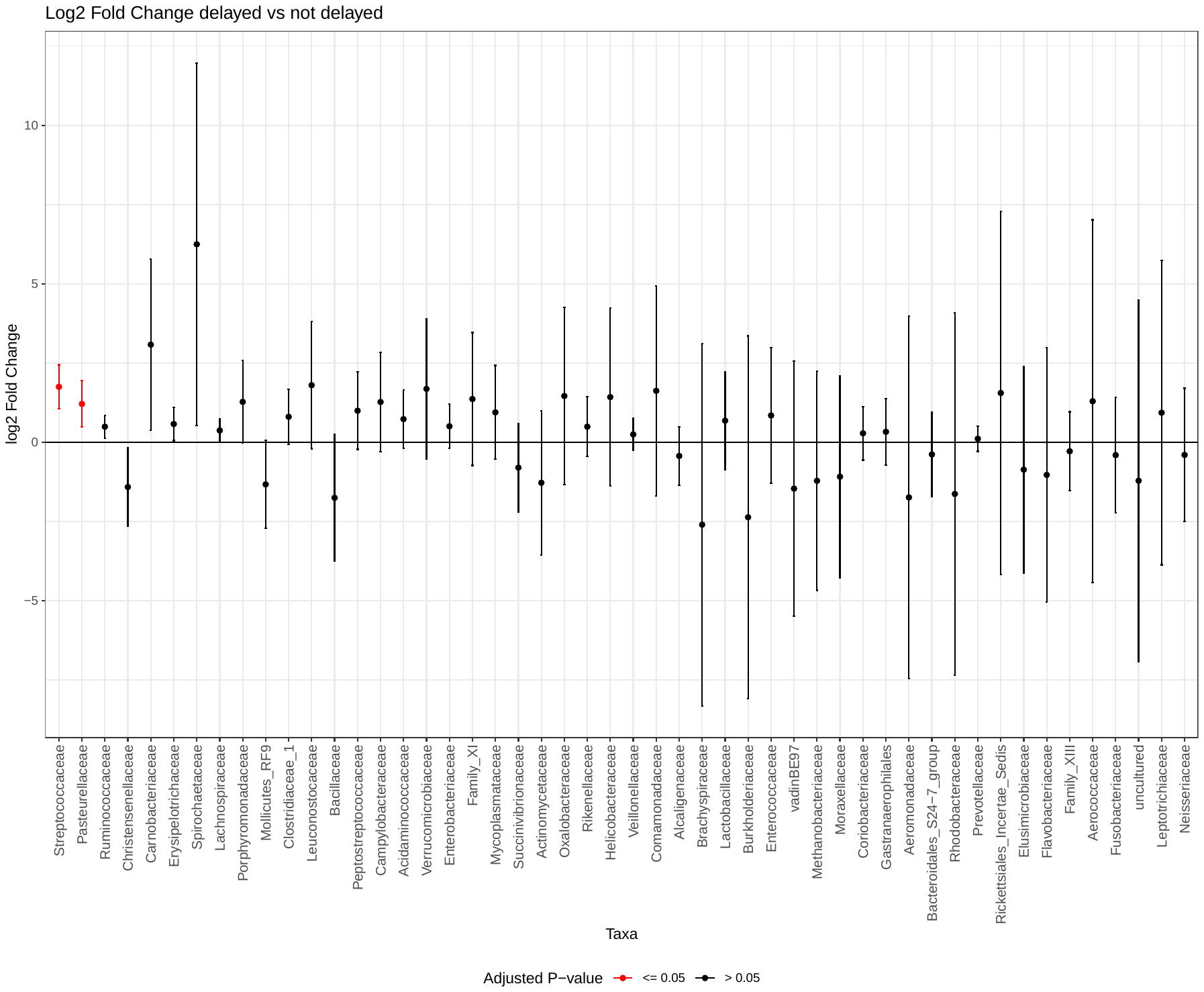

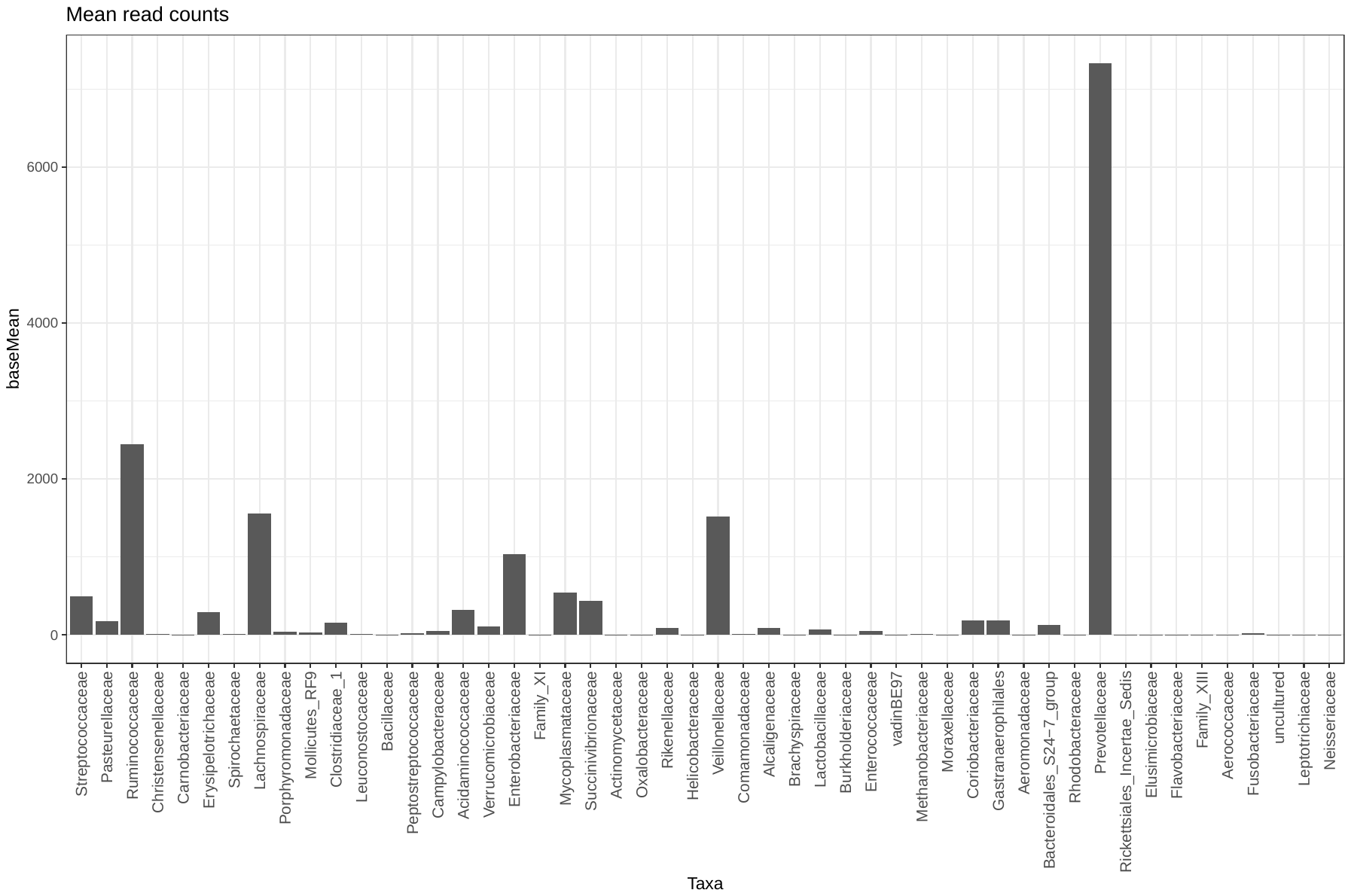

**SUPPL FIGURE 4.** Results of the association between the developmental overall score and individual bacterial species using DESeq2 in the lowest and highest quartiles of the score distribution. For the overall score: A) Mean base counts for the first 50 families ordered according to the adjusted p-value. B) log2 fold change for the first 50 families ordered according to the adjusted p-value. For the Problem-Solving score: C) Mean base counts for the first 50 families ordered according to the adjusted p-value. D) log2 fold change for the first 50 families ordered according to the adjusted p-value.

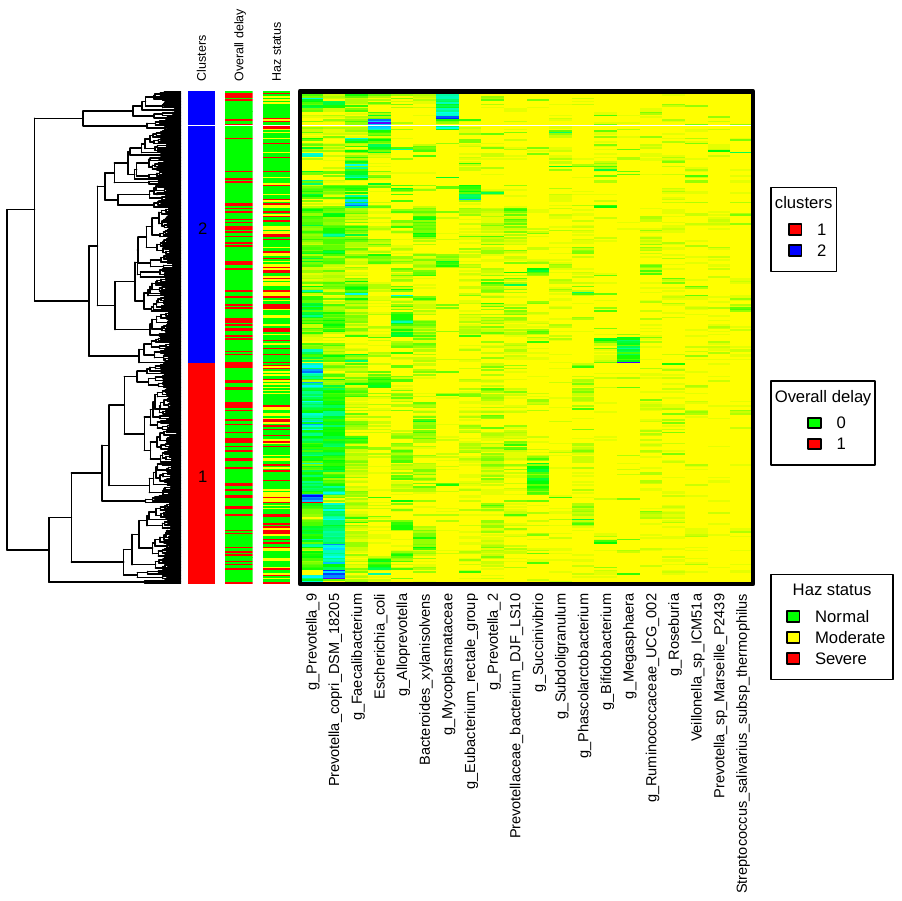

**SUPPL FIGURE 5.** Heatmap of the 20 most abundant bacterial amplicon sequencing variants’ relative abundance after ascending agglomerative hierarchical clustering of participants based on the Bray-Curtis distance and Ward linkage. The hierarchical clustering resulted in two community clusters (1 and 2) used in subsequent analyses.

HAZ: Height-for-Age Z-score.
